# Risk and early signs of PTSD in people indirectly exposed to October 7 events

**DOI:** 10.1101/2023.12.15.23300048

**Authors:** Dan Yamin, Shahar Lev-Ari, Merav Mofaz, Ron Elias, David Spiegel, Matan Yechezkel, Margaret L. Brandeau, Erez Shmueli

**Author notes:** Contributed equally (Dan Yamin, Shahar Lev-Ari, Erez Shmueli). **Address correspondence to**: Prof. Erez Shmueli. **Primary Funding source:** ISRAEL SCIENCE FOUNDATION (grant no. 3409/19), within the Israel Precision Medicine Partnership program, and Koret Foundation gift for Smart Cities and Digital Living.

## Abstract

The coordinated terrorist attacks on October 7, 2023, resulted in catastrophic atrocities, and marked the beginning of the 2023 Israel–Hamas war. The overwhelming coverage by mainstream and social media, characterized by extreme details and graphic images, vividly transported viewers to the horrifying scene. It remains unclear to what extent such indirect exposure influences the occurrence of stress, anxiety, and post-traumatic symptoms. We analyzed data from a three-year prospective study in which 4,797 participants received smartwatches and completed daily questionnaires, supplemented by a nationwide clinical survey with 2,536 participants. Among the participants not directly exposed, we estimated PTSD prevalence to be 22.9-36.0% and moderate to severe anxiety prevalence to be 22.9-55.32%, with 752,057 daily questionnaires before and after October 7 further indicating higher stress levels than those reported in previous events, including political disputes, the COVID-19 pandemic, and past armed conflicts. The occurrences of PTSD and anxiety are well explained by increased and persistent news consumption, and especially by the availability of gory videos on social media. Continuous monitoring of participants via smartwatches and daily questionnaires further revealed considerable differences in stress, mood, step counts, sleep quality, and duration in the first week after the October 7 events among those who later developed PTSD. This study demonstrates the unprecedented amplifying effect of mass media on mental health in terror and war settings and highlights the potential of continuous monitoring for early detection and prompt treatment of those in need.

## Main text

On October 7, 2023, a series of attacks across Israeli municipalities resulted in significant casualties, sexual abuse, and abductions of civilians from their homes ^1^. These atrocities not only had a visible toll, including physical destruction and loss of life, but also raised concerns about potential long-term mental health implications, such as post-traumatic stress disorder (PTSD) among civilians.

Early detection of PTSD is crucial, as timely interventions can significantly aid individuals in coping with the initial impact and prevent the progression to chronic PTSD. Beyond certain preconditions ^2^, two primary factors influence an individual’s risk of developing PTSD: the extent of their exposure to the trauma and their unique physiological and psychological responses in the aftermath ^2^. Previous research indicates that indirect exposure to trauma, such as through media, can also precipitate PTSD, although the effects may differ from those of direct exposure ^3–5^. The graphic coverage of the October 7 events on social media platforms like Telegram and TikTok was extremely vivid and detailed, posing new challenges in assessing the media’s impact on PTSD. In terms of individual responses to trauma, the ‘impact phase’—the initial period post-trauma—is critical in the development of PTSD. This phase involves the initial recognition and reaction to the traumatic event, though not everyone who experiences this stage will necessarily develop chronic PTSD. The relationship between individuals’ responses during this stage and the risk of developing PTSD at later stages remains an area requiring further exploration.

Stress response systems, both the sympathetic adrenal medullary system producing epinephrine and rapid increases in heart rate and blood pressure, and the hypothalamic- pituitary-adrenal axis releasing increased cortisol that mobilizes blood glucose, are designed for rapid response to stressors, with homeostatic mechanisms that return to relatively normal levels quickly after stress is over. However, chronic or repeated stress produces a resetting of response levels, termed allostasis, a new and debilitating neuroendocrine pattern ^6^.

Neuroplasticity, especially in portions of the brain devoted to memory and emotion such as the dentate gyrus of the hippocampus, strengthens pathways that trigger ongoing physiological stress response, including hyperarousal at reminders of earlier or ongoing stressors, laying the neural groundwork for PTSD ^7^. Prior prospective studies have shown an evolution of patterns of PTSD symptoms over time. A network analysis of populations exposed to trauma ^8^ found initial patterns of reexperiencing trauma, intrusive thoughts, and avoidance, while a year later avoidance was associated with dysphoria, and emotional numbing was connected with social distancing ^8^. A study of responses to the 9/11 World Trade Center attack found that early emotional support and lack of constraints on emotional expression as well as lack of cognitive negativity and self-blame predicted less emotional distress six months later. Media exposure was associated with substantially higher initial distress ^9^. These findings all point to possible avenues of management of response to community trauma that may ameliorate emotional damage over time.

To bridge these gaps and gain insights into the early indicators of PTSD risk, our analysis encompassed data from a three-year observational prospective study of 4,797 participants. These individuals were provided with smartwatches and tasked with completing daily questionnaires using a dedicated mobile application, capturing data both before and following the events of October 7. Complementing this, we conducted a panel study including nationwide surveys involving 2,536 participants, ensuring a representative sample of the adult Jewish population in Israel.

### Unprecedented levels of stress, anxiety, and PTSD

On the morning of October 7, during a time usually marked by holiday calm and weekend relaxation, a series of unexpected atrocities jolted the Israeli population, triggering unprecedented levels of stress, anxiety, and post-trauma. The extent of this upheaval was captured through detailed data from daily self-reported questionnaires and a variety of physiological metrics measured by smartwatches worn by participants in our prospective study (Figure 1 A-E). Specifically, over the past three years, we conducted an observational prospective study in which we equipped 4,797 participants with smartwatches and a dedicated mobile application ^10–14^. The smartwatches continuously monitored several physiological measures, while the mobile application collected daily self-reported questionnaires on the general health and well-being of each participant. Data based on 752,057 daily questionnaires filled out by participants between January 1, 2021 and November 30, 2023 revealed that stress levels after October 7 were unprecedented (a maximum level of 3.45, 95% CI 3.3-3.6, on a scale of 1 to 5; ‘very low’– ‘very high’) and far exceeded those observed during previous events (Figure 1A). This includes the deadliest and most contagious COVID-19 wave, during which 30% of the Israeli population tested positive over a short period of three months (a maximum level of 2.44, 95% CI 2.4-2.48); widespread political protests sparked by a controversial judicial reform, culminating in the dismissal of an Israeli minister and strikes across private and public sectors (a maximum level of 2.6, 95% CI 2.54-2.66); and intense military engagements, with over 1,000 rockets launched from Gaza towards multiple Israeli cities (a maximum level of 2.94, 95% CI 2.83-3.05). Additionally, data comparison of a routine period preceding October 7 (September 30, 2023 until October 6, 2023) and the week after October 7 highlighted a marked decline in reported mood level (3.68, 95% CI 3.59-3.77 vs. 2.81, 95% CI 2.69-2.93, on a scale of 1 to 5); decreased physical activity, as evidenced by a significant drop in daily step count (7,881, 95% CI 7,588-8,175 vs. 6,127, 95% CI 5,841-6,411 steps); and a distinct decrease in reported sleep quality (3.51, 95% CI 3.41-3.6 vs. 3.14, 95% CI 3.03-3.25, on a scale of 1 to 5) accompanied by an increase in awake time during the night (594.25, 95% CI 544.84-643.66 vs. 687.35, 95% CI 628.82-745.88 seconds) (Figure 1B-E). As an illustrative demonstration, we found increase in heart rate and stress based on heart rate variability during the first 72 hours after October 7 (Supplementary Figures S2 and S3) that were comparable to the reactions we previously observed following COVID-19 vaccinations ^11^.

**Figure 1:**
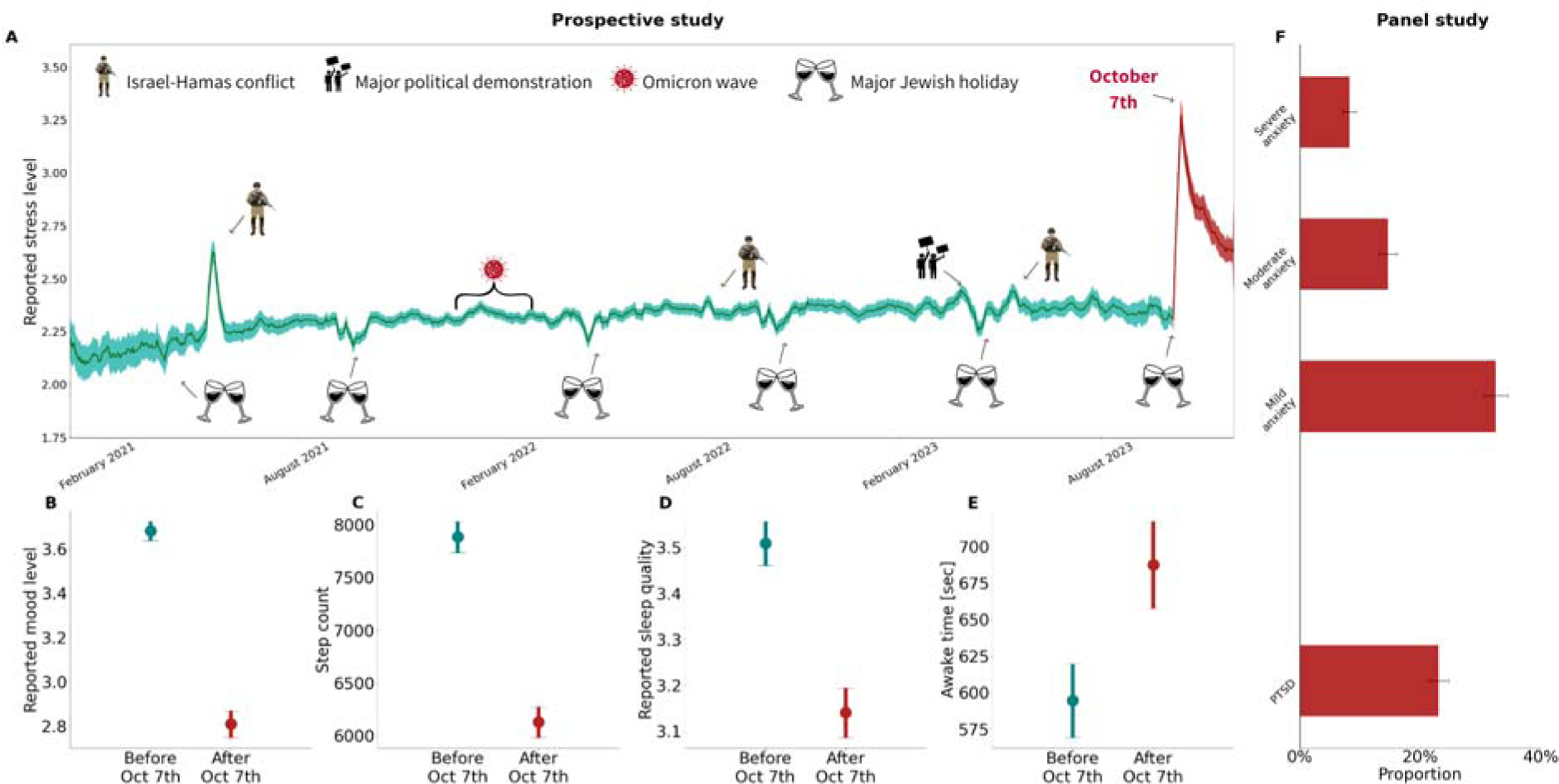
(A) mean daily reported stress levels for the prospective study active participants for the period January 1, 2021, to November 30, 2023. The stress levels were reported by participants as part of 752,057 daily questionnaires filled out. Stress was reported on a scale of 1 (Low) - 5 (High). The presented values are the daily moving average and the associated 95% confidence interval using a 7-day window. Figures (B-E) compare the measures recorded in a routine period before October 7 (September 30, 2023, to October 6, 2023) with those from the week after. Mean values and 95% confidence intervals are presented. Figure F shows the prevalence estimates of anxiety levels using the GAD-7 questionnaire and PTSD using the PCL-5 questionnaire among the panel study participants who were indirectly exposed to the October 7 events. In all panels, the color red represents data recorded after October 7, and green represents data recorded before October 7.

Next, we evaluated the extent to which the atrocities had post-traumatic effects on those indirectly exposed (Figure 1F). To do so, we conducted a Post-traumatic Stress Disorder Checklist (PCL-5) survey among a panel of 2,536 participants 6-7 weeks after October 7. The PCL-5 questionnaire has previously demonstrated strong internal consistency, high test-retest reliability, and strong convergent validity with the gold standard PTSD diagnosis ^15^. Participants were representative of the adult Jewish population in Israel by age, sex, and geographical location. Among the 2,341 participants who were indirectly exposed to the atrocities, the estimated prevalence of PTSD was 22.9% (95% CI 21.19%-24.6%) using the most conservative case definition, and up to 35.96% (95% CI 34.05%-37.93%) with the least conservative definition. This prevalence was substantially higher than the 7.5% found 6-8 weeks after the 9/11 terror attack among New York City residents ^16,17^.

Similarly, using the General Anxiety Disorder (GAD) 7-item questionnaire among those not directly exposed, the estimated prevalence of anxiety was 55.32% (95% CI 53.23%-57.4%), with 22.86% (95% CI 21.14%-24.63%) being moderate to severe.

### Exposure to news and gory videos as predictors of the prevalence of PTSD

In contrast to the 9/11 terror attack, where smartphones and social media platforms were not available, the October 7 events were extensively documented in real-time through GoPro cameras and disseminated on various media platforms. We investigated the relationship between the frequency of news consumption and the number of gory videos viewed, and the prevalence of PTSD among panel participants (Figure 2). After adjusting for age, sex, socioeconomic status, and religious level, logistic regression revealed a positive correlation: higher news consumption in the first week and more gory videos viewed following October 7 were associated with an increased prevalence of PTSD (p.values 0.041 and 0.003) (Supplementary Table S3). For instance, estimated PTSD prevalence among those who did not watch any gory videos or news was 7.7%, compared to 31.7 % among those who watched more than eight hours of news per day and viewed five or more gory videos during the first week after the event (Figure 2A). Among the various media platforms, Telegram presented the highest exposure rate to gory videos at 88.4%, followed by Instagram and TikTok at 59.3% and 55.4%, respectively (Figure 2B).

**Figure 2:**
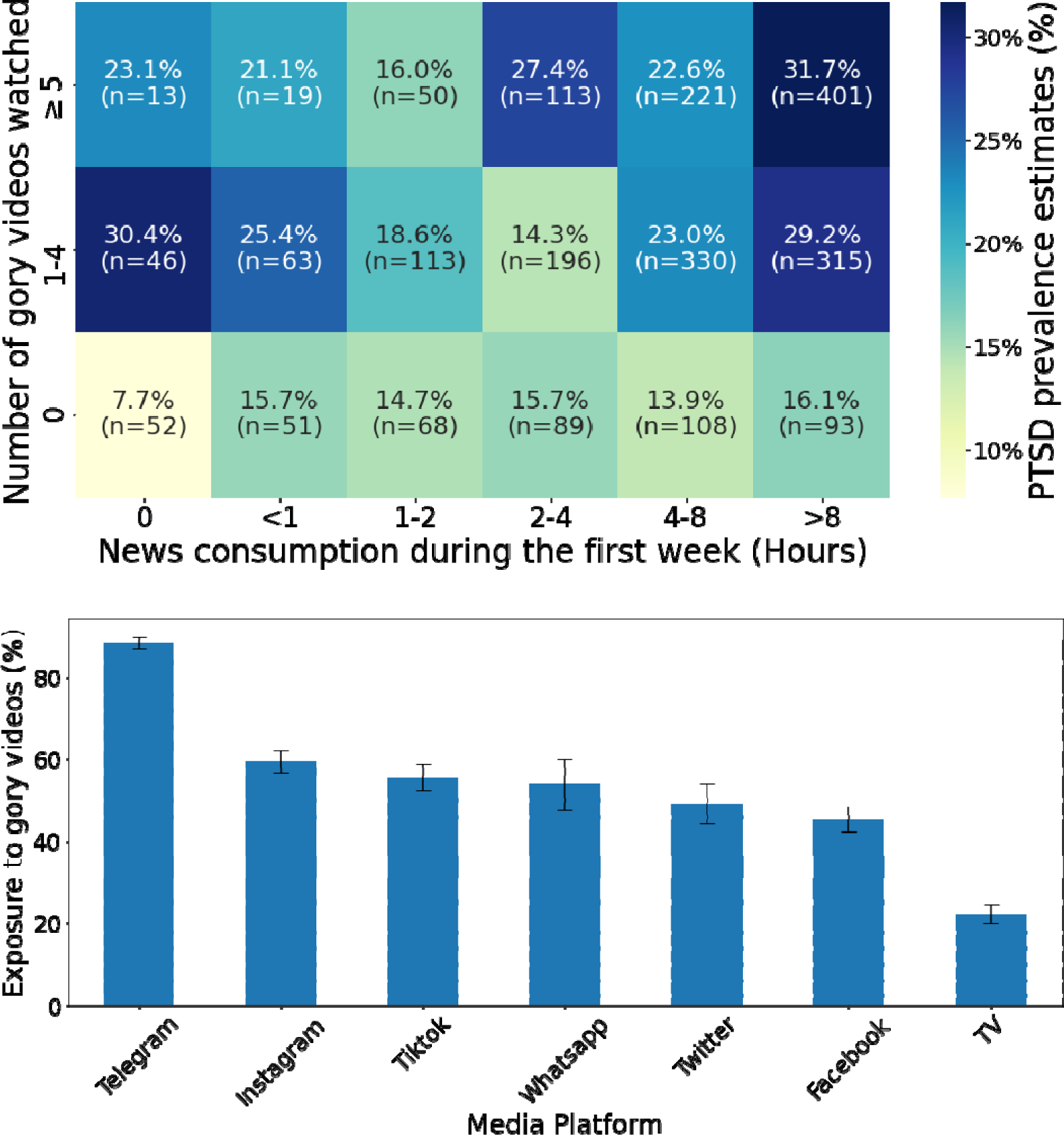
(A) Estimated prevalence of PTSD as a function of the number of hours spent watching news each day and the number of gory videos watched, in the first week after October 7. Adjusting for age, sex, socioeconomic status, and religious level, a logistic regression confirms that ‘news consumption’ and ‘number of gory videos watched’ are significantly correlated with PTSD prevalence (p.value 0.041 and 0.003 respectively) (Supplementary Table S3). (B) Exposure rate to gory videos by media platforms. For each media platform, we calculated the exposure rate as the number of participants who reported watching gory videos in that media platform divided by the total number of individuals who reported using that media platform.

### Greater changes in measures among those who will later develop PTSD

Interventions administered shortly after a traumatic event have the potential to prevent post- traumatic stress disorder ^18^. A key challenge in delivering interventions is understanding how PTSD develops in the acute post-trauma period ^18^. Specifically, little is known about physiological reactions occurring shortly after the trauma for those who later develop PTSD ^19^. Focusing on the first week after the events, we found that participants who later developed PTSD exhibited a higher increase in reported stress levels (1.31 vs. 0.85, on a 1-5 scale) and a sharper decline in: reported mood level (1.28 vs. 0.82, on a 1-5 scale), physical activity as manifested by step count (2155 vs. 1620 steps), reported sleep quality (0.7 vs. 0.31, on a 1-5 scale), and reported sleep duration (31 vs. 10 minutes) (Figure 3).

**Figure 3:**
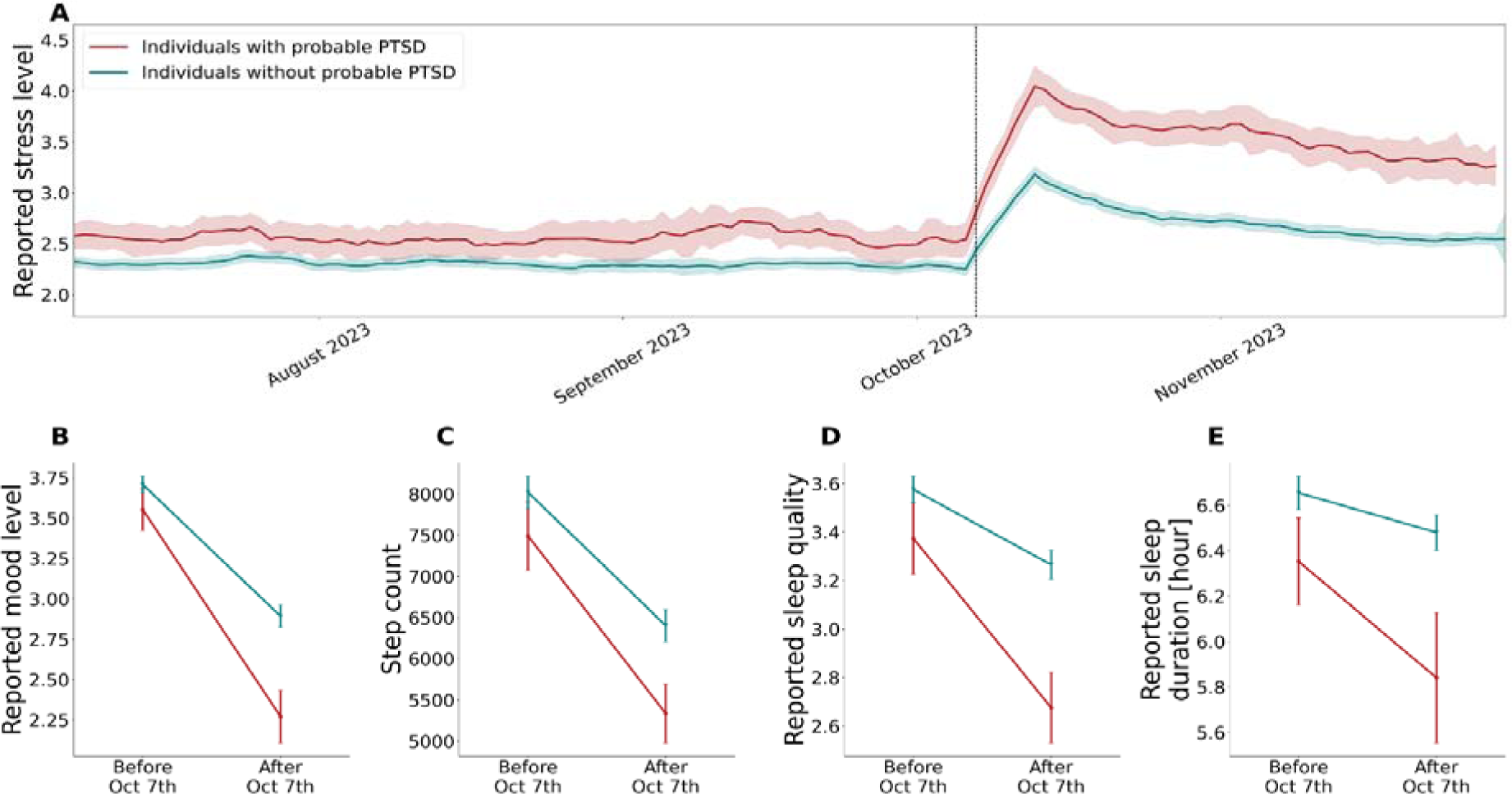
(A) Mean daily reported stress level for the prospective study participants for the period July 1, 2023 to November 30, 2023, stratified by PTSD status. Stress levels were reported by participants as part of the daily questionnaires on a scale of 1 (Low) - 5 (High). The presented values are the daily moving average and the associated 95% confidence interval using a 7-day window. Figures (B-E) compare the measures recorded in a routine period before October 7 (September 30, 2023, to October 6, 2023) with those from the week after, stratified by PTSD status. Mean values and 95% confidence intervals are presented.

**Figure 4:**
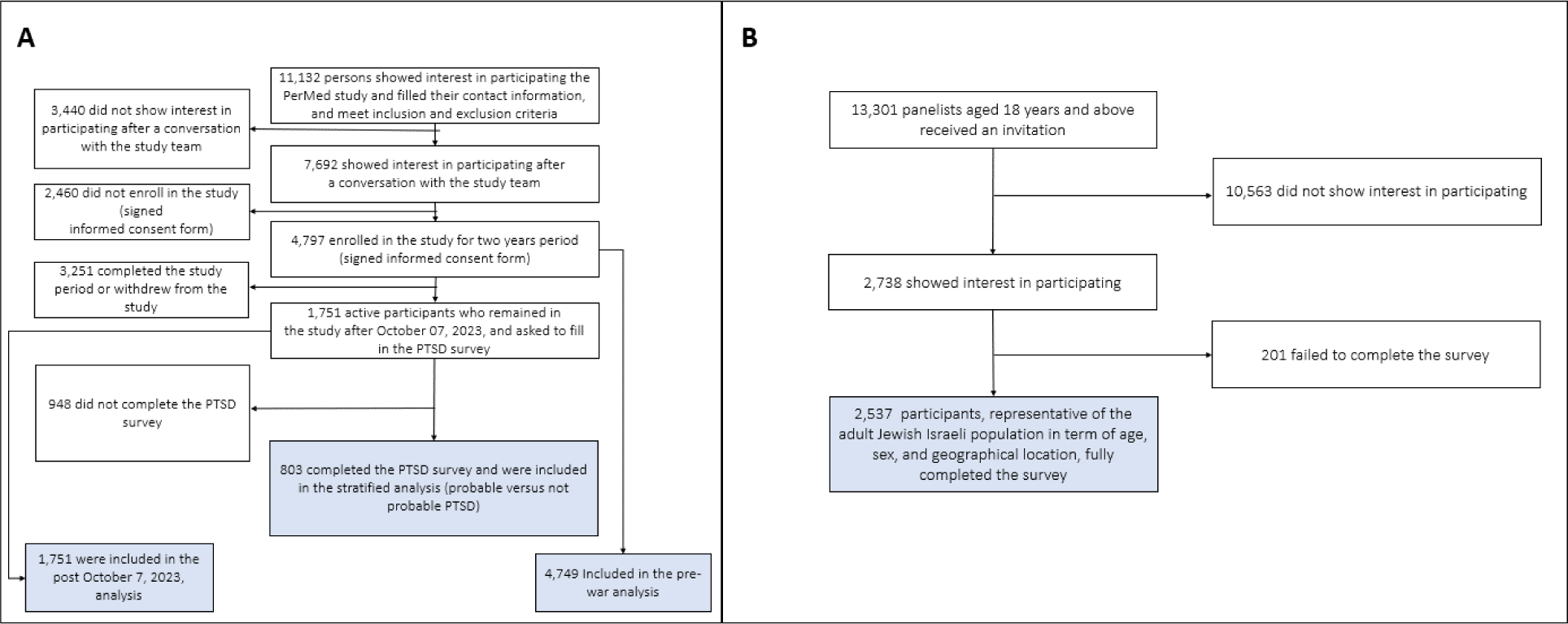
Study participants. (A) prospective study, (B) panel study.

## Discussion

In today’s reality, where detailed, unfiltered, graphic, and prolonged coverage of massive traumatic events is common, we find that PTSD rates among those indirectly exposed can be substantially high, reaching 22.89-35.96%. These rates are exceptionally high, comparable to those experienced by health professionals who encounter trauma in their professional duties ^20–22^. Specifically, the hours spent, and the number of gory videos viewed in the first week following October 7 were associated with an increased risk. Furthermore, using information from smartwatches and simple daily questionnaires, we found early signs in individuals who later developed PTSD: these include a substantial increase in stress level and decline in mood level, reduced activity as indicated by decreased step counts, and abnormal sleep as reflected by a decline in sleep quality and duration.

The DSM-5’s inclusion of indirect exposure to trauma as a criterion for PTSD acknowledges the reality of ‘secondary traumatic stress’, a condition prevalent among professionals regularly exposed to the aftermath of traumatic events, such as first responders and mental health workers ^23^. Highly visual social media platforms, including Telegram, Instagram, and TikTok, are gaining popularity, particularly among younger demographics ^24^. Our study emphasizes the profound impact that mass media’s graphic portrayals of trauma can have on public mental health in terror and war contexts, highlighting the public health importance of developing guidelines and recommendations for posting and watching such unfiltered content. Given the ubiquity of video and still photography on smartphones and other portable cameras, people are now widely exposed to real images and sounds of violence, murder, sexual assault, and other trauma as it occurs. Such exposure produces post-traumatic stress symptoms that are comparable to those that follow current Criterion A exposure ^25^, prompting a need for consideration of further expansion of exposure criteria in the DSM to include seeing such images electronically and outside the context of work.

Few studies have explored the potential of physiological data from smartwatches in assessing individual responses during the immediate aftermath of trauma. Recent study found that monitoring such biometrics could predict adverse posttraumatic neuropsychiatric sequelae following traumatic stress exposure in hospital settings ^26^. Another study demonstrated the efficacy of detecting hyperarousal through heart rate and body acceleration data, achieving an 83% success rate in predicting PTSD onset in veterans ^27^. Our study expands this knowledge by revealing that civilians indirectly exposed to secondary traumatic stress via mass media suffer from similar types of physiological changes in stress, sleep, and physical activity during the week after such exposure and are at increased risk of developing PTSD.

Our findings extend current knowledge linking heightened stress levels, decreased physical activity, and sleep disturbances to the development of PTSD ^28–33^. Further, our findings suggest that the extent of these physiological reactions, as detected objectively via smartwatches and subjectively by the individual, provides early signs from the impact phase. The significance of these findings is amplified by the fact that a substantial number of individuals with PTSD remain undiagnosed, despite the availability of early and personalized treatments ^34–38^. The absence of proper diagnosis and subsequent treatments can lead to more severe long-term health outcomes, including persistent depressive symptoms, increased suicide attempts, higher morbidity, and premature death^38–40^. The ability to monitor and analyze biometric data from smartwatches opens new avenues for identifying early signs of PTSD at the population level, particularly in cases of indirect exposure to trauma through mass media. This is crucial for timely intervention and the prevention of long-term adverse mental health outcomes.

Our study has several limitations. We assumed that the traumatic event is defined by the October 7 atrocities, but the context of our PTSD prevalence estimates includes the onset of war following the October 7 events in the Gaza Strip. However, as opposed to the ongoing trauma of the civilians in Gaza, we believe that the trauma experienced by Israeli citizens primarily stems from the specific incidents on October 7. Second, though previously validated in several settings, including in Israel ^15,41^, our prevalence estimate for PTSD is based on the PCL-5 survey, while a gold standard diagnosis requires assessment by healthcare professionals. Finally, the lack of longitudinal data limits our understanding of PTSD’s progression, severity, and long-term impacts. Future research should include longer follow-up periods to provide a more comprehensive understanding of the trajectory of PTSD over time.

## Acknowledgments

This work was supported by ISRAEL SCIENCE FOUNDATION (grant no. 3409/19), within the Israel Precision Medicine Partnership program and a Koret Foundation gift for Smart Cities and Digital Living.

## Authors’ contributions

Conception and design: DY, SLA, ES. DY, MY, MF, RE, and ES had access to the raw data and were responsible for verifying the data. Analysis and interpretation of the data: MY, DY, ES, MF, RE, and DS. Statistical analysis: MY, DY, ES, MF, and RE. Drafting the article: all authors. Critical revision of the article for important intellectual content: DY, ES. Final approval of the article: All authors. Obtaining funding: DY, ES, and MLB.

## Methods

### Study Design and Participants

#### Panel Study

For the panel study, we hired a professional survey company to recruit 2,536 participants from November 1, 2023 to December 3, 2023. Participants were aged 18 years or older, and were representative of the adult Israeli Jewish population by age, sex, and geographical location (Supplementary appendix A). Participant recruitment was conducted through a preregistered panelists’ database via email. The survey company was responsible for ensuring that participants fulfilled all the study’s requirements. To guarantee the representativeness of the sample, invitations were systematically distributed to panelists, ensuring their participation in the study through a staggered and quota-based approach. For inclusion in the panel study, participants were required to be 18 years or older, enrolled in the survey company’s database of panelists, and willing to consent to the specified terms of the study (Supplementary appendix D). Panelists were excluded if they failed to complete the study requirements.

#### Prospective study

The ongoing PerMed prospective observational study included 4,797 participants aged 18 years and older who were recruited starting November 1, 2020, from all across Israel ^1–5^(Supplementary appendix A and B). As of December 6, 2023, a total of 1,751 participants remained active in the study. Participant recruitment was conducted via advertisements on social media and word-of-mouth. Each participant signed an informed consent form after receiving a comprehensive explanation of the study from a professional survey company.

To be included in the PerMed study, participants needed to be 18 years or older, using their own smartphone, and able to give written informed consent by themselves. Participants were excluded if they knew that they would be outside of Israel for more than three months continuously at any point for the two years after enrolment. Participants were also excluded if they were students or employees of the principal investigator.

### Procedures

#### Panel study

Upon enrollment, participants were provided with written information about the nature of the study. We asked participants to complete online PTSD and anxiety surveys, focusing on coping with events that occurred on October 7, to detect probable cases of PTSD and anxiety at the nationwide level. At this stage, participants were also requested to provide demographic information, including age, sex, household size, educational background, and news consumption behavior.

The online PTSD survey incorporates the Post-traumatic Stress Disorder Checklist (PCL-5), a self-report questionnaire designed to assess the 20 symptoms outlined in the DSM-5 criteria for PTSD. The PCL-5 serves various purposes, including screening individuals for PTSD and making provisional diagnoses ^6^. It is widely used for clinical and research purposes; psychometrically, the PCL-5 demonstrates strong internal consistency (α = .94), high test- retest reliability (r = .82), and strong convergent validity (rs = .74 to .85) ^6^.

The survey also utilized the General Anxiety Disorder (GAD) 7 item questionnaire (GAD-7) to identify probable cases of GAD along with measuring anxiety symptom severity. The tool is also widely used as a screening measure of panic, social anxiety, and PTSD. The questionnaire is considered valid, sensitive, and specific for the diagnosis of GAD in the general population ^7^. We used the Israeli Ministry of Health’s (IMOH) Hebrew translation of these two questionnaires.

#### Prospective study

Upon enrolment in the prospective study, we collected information on participants’ sex, age, household income, city of residence, and religious level. After enrolment, participants used the PerMed mobile application to complete a daily questionnaire ^8–10^. The questionnaire allowed participants to report on stress levels, mood, and sleep quality (Supplementary appendix A). Participants who were active in the study after the events on October 7, 2023, were asked to fill in an online PTSD survey eight weeks later.

Participants in the prospective cohort were equipped with Garmin Vivosmart 4 smart fitness trackers. Among other features, the smartwatch collects data for all-day heart rate, HRV- based stress, ^11^ step counts, sleep duration, and sleep-level classification, including light, deep, rapid eye movement (REM), and awake periods ^1^ (Supplementary appendix A). We focused on these measures because they provide continuous information on the well-being of an individual. Additionally, individuals diagnosed with PTSD may encounter nightmares, insomnia and often report a reduction in sleep time due to waking up earlier than desired, restlessness during the night, and difficulties falling asleep, all of which impact daily life ^12^. Step counts are strongly correlated with the physical activity level of an individual. Given that PTSD involves anxiety and depression, those diagnosed with PTSD tend to be less physically active ^13^.

The smartwatch’s optical wrist heart rate monitor was designed to continuously measure the user’s heart rate. The frequency at which the heart rate was measured varied and sometimes depended on the level of activity of the user: when the user started an activity, the optical heart rate monitor’s measurement frequency increased. As heart rate variability (HRV) was not easily accessible through Garmin’s application programming interface, we used Garmin’s stress level instead, which is calculated based on HRV ^11^. Specifically, the device uses heart rate data to determine the interval between each heartbeat. The variable length of time between each heartbeat is regulated by the body’s autonomic nervous system. Less variability between beats correlates with higher stress levels, whereas an increase in variability indicates less stress ^14^. When examining the data collected in our study, we identified a heart rate sample approximately every 15 s and a stress-based HRV sample every 180 s.

We developed a dedicated data collection platform that collects for each participant data from the smartphone sensors and daily questionnaires via the PerMed application and the smartwatch sensors via the Garmin Health API (Supplementary appendix B). These collected data are securely stored in Tel Aviv University facilities.

Participants completed a one-time enrolment questionnaire, received Garmin Vivosmart 4 smartwatches, and installed two applications on their mobile phones: (1) the PerMed application ^8–10^, which collects daily self-reported questionnaires; and (2) an application that passively records smartwatch data. Participants were asked to wear their smartwatches as much as possible. Using a dedicated dashboard to monitor compliance, a survey company ensured that participants’ questionnaires were filled at least twice a week, that their smartwatches were charged and properly worn, and that any technical problems with the mobile applications or smartwatch were resolved (Supplementary appendix A).

We implemented several preventive measures to minimize participant attrition and discomfort and thereby improve the quality, continuity, and reliability of the collected data. First, each day, participants who did not fill out their daily questionnaire by 19:00h received a reminder notification through the PerMed application. Second, we developed a dedicated dashboard that allowed the survey company to identify participants who repeatedly neglected to complete the daily questionnaire or did not wear their smartwatch for extended periods of time. These participants were contacted by the survey company (either by text message or phone call) and were encouraged to better adhere to the study protocol. Third, to strengthen participants’ engagement, a weekly summary report was generated for each participant, which was available inside the PerMed application. Similarly, a monthly newsletter with recent findings from published studies and useful tips regarding the smartwatch’s capabilities was sent to the participants. At the end of two years, participants received all the obtained personal insights and could keep the smartwatch as a gift (Supplementary appendix C).

### Outcomes

For both the prospective and panel study cohorts, we analyzed the online PTSD survey (Supplementary Table S1 and Supplementary appendix D). We defined the main outcomes as probable PTSD and anxiety levels for each participant.

For the prospective study, we analyzed step counts, sleep duration, and awake episodes during sleep from the smartwatches. In analyzing the daily questionnaires, we calculated the daily mean of reported stress, reported mood, reported sleep quality, and reported sleep duration before and after the events of October 7. For each measure, the prespecified outcomes are the daily mean value of each measurement. We calculated the daily mean value before and after the events of October 7.

We also analyzed heart rate and HRV-based stress from the smartwatches. For these measures, we calculated the hourly mean value before and after the events of October 7.

### Statistical analysis

PTSD diagnosis requires an interview conducted by a trained clinician. As our outcome was derived from a screening tool, we characterize the result as “probable PTSD”. We defined probable PTSD as a total score of 33 on the PCL-5 questionnaire and meeting the DSM-5 diagnostic rule which requires at least: 1 B item (questions 1-5), 1 C item (questions 6-7), 2 D items (questions 8-14), 2 E items (questions 15-20). This definition aligns with the most conservative criteria, consistent with the guidelines established by the International Society for Traumatic Stress Studies ^15^ (Supplementary appendix D). In a secondary analysis, we considered a less conservative definition of probable PTSD, whereby it is identified by either a total score of 31 on the PCL-5 questionnaire or by meeting the DSM-5 diagnostic rule.

In line with the GAD-7 criteria, the anxiety level of participants was determined by the total score: no anxiety (0-4), mild (5-9), moderate (10-14) and severe (15) ^7^.

#### Panel study

##### Descriptive statistics

We analyzed the data of participants who had indirect exposure to the events of October 7. Specifically, we defined indirect exposure for all participants who, along with their immediate family, were not injured, killed, or abducted. To ensure specificity in attributing PTSD sources, we excluded individuals who evacuated their homes. This exclusion is based on the consideration that PTSD in these cases may stem more from the evacuation process than the events of October 7. We evaluated the estimated prevalence of PTSD as the proportion of individuals with probable PTSD. We also stratified the prevalence estimate by the duration of news information consumption during the first week after October 7 and the extent of exposure to gory videos. Additionally, we examined the rates of each anxiety level.

We also evaluated the rates of exposure to gory videos on each media platform. Specifically, we divided the number of participants who were exposed to gory videos on each platform by the number of participants who consumed news information using that platform.

The 95% confidence intervals (CIs) were obtained under the assumption of binomial distribution.

##### Inferential statistics

To examine the unadjusted associations between the probability of probable PTSD, duration of news consumption during the first week following October 7, and the extent of exposure to gory videos, while controlling for other explanatory variables (age, sex, educational background, PTSD background, religious level, and socioeconomic level), we applied a logistic regression model:

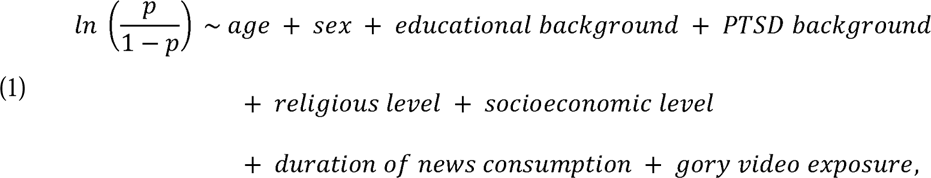

where p represents the probability of probable PTSD. Age is treated as a continuous variable, while educational background, religious level, socioeconomic level, duration of news consumption, and gory video exposure are considered ordinal variables. Sex and PTSD background are categorized as Boolean variables.

Likewise, to examine the unadjusted associations between the probability of moderate to severe anxiety, duration of news consumption during the two weeks before filling in the online PTSD survey, and the extent of exposure to gory videos, while controlling for other explanatory variables (age, sex, educational background, anxiety background, religious level, and socioeconomic level), we applied a logistic regression model:

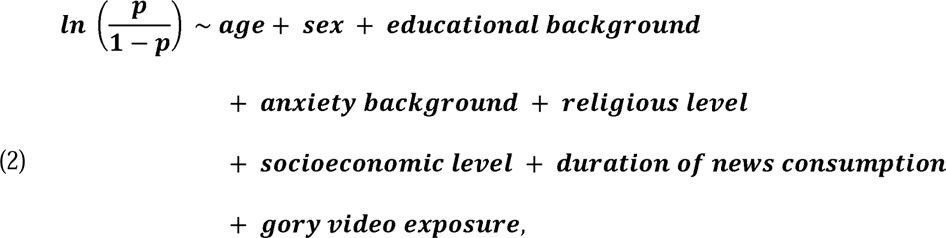

where p represents the probability of moderate to severe anxiety. Age is treated as a continuous variable, while educational background, religious level, socioeconomic level, duration of news consumption, and gory video exposure are considered ordinal variables. Sex and anxiety background are categorized as Boolean variables.

For both models, we derived the odds ratios and their corresponding 95% confidence intervals from the values of the regression coefficients.

#### Prospective study

##### Descriptive statistics

From the commencement of the prospective study on January 1, 2021, to December 3, 2023, we collected 1,933,475 days of smartwatch data and 752,057 completed self-reported questionnaires. Additionally, 803 participants filled in the PTSD online survey.

Before analyzing the data, we conducted a preprocessing step. If participants filled out the daily questionnaire more than once on the same day, only the latest questionnaire for that day was considered. The rationale behind this decision was that once a questionnaire was filled, it was sent to the server, and could not be updated. Therefore, in the case of a filling error, participants were instructed to re-fill the questionnaire.

For a more comprehensive understanding of individual perception, we examined the reported stress levels throughout the PerMed study period. The reported stress levels were recorded from the daily questionnaires on a scale of 1 to 5, where 1 represents ‘very low,’ and 5 signifies ‘very high.’ We calculated the daily moving average and the associated 95% confidence interval with a 7-day window from January 1, 2021, to November 30, 2023, for all active participants. For participants who filled in the PTSD online survey, we implemented the same procedure, stratifying them based on whether they exhibited probable PTSD.

We noted several significant national events that might have influenced reported stress levels, including the major Jewish holidays (Passover and Rosh Hashanah), armed conflicts involving the firing of at least 1,000 rockets at the civilian population, a major political demonstration related to judicial reform that was escalated on March 26, 2023, following the dismissal of the Minister of Defense and the strike in both the public and private sectors, and the SARS-CoV-2 Omicron wave, which persisted from December 20, 2023, to May 1, 2023, with approximately 30% of the Israeli population reportedly infected ^16^.

For each participant who remained active after October 7, 2023, we assessed the reported sleep quality (measured from daily questionnaires on a scale of 1 to 5 where 1 represents ‘awful’ and 5 signifies ‘excellent’), as well as various physiological indicators: duration of awake time during sleep, step counts, and duration of high-stress levels. We computed a single weighted average value for each of the two periods—the week prior to October 7, 2023 (baseline period) and the week after October 7, 2023.

The weighted average for each participant was determined by first averaging the corresponding indicator values separately for workdays and free days (weekends and national holidays). Subsequently, the weighted average of these two values was calculated, assigning a weight of 5/7 to workdays and 2/7 to free days. The rationale for computing this weighted average is further explained elsewhere ^1^. In essence, we identified a weekly rhythm across various indicators, with free days exhibiting different mean daily values than workdays. Given the relatively short duration of the examined time periods and variations in the number of free days (e.g., holidays), we aimed to correct for potential biases. Finally, we calculated the mean and corresponding 95% confidence interval for each indicator across the participants. For each indicator, we utilized a paired t-test (Supplementary Table S5) to examine the difference between the weighted averages during the baseline period and those following October 7, 2023.

For participants who filled in the PTSD online survey, we repeated the above analysis, stratifying participants based on whether they exhibited probable PTSD.

##### Inferential statistics

For each of the well-being indicators, we explored the unadjusted associations between the probability of probable PTSD, the weighted average during the baseline period, and the difference in the weighted average between baseline and the week after October 7 periods, while controlling for other explanatory variables (age and sex), we applied a logistic regression model (Supplementary Table S6):

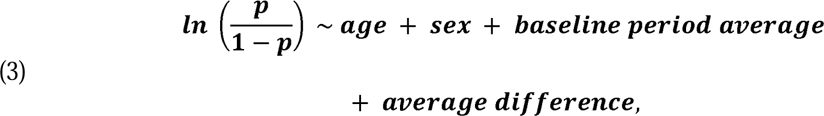

where p represents the probability of probable PTSD. Age, weighted average during the baseline period, and the difference in the weighted average between baseline and the week after October 7 periods are treated as continuous variables, while sex is categorized as Boolean variables.

All statistical analyses were performed using Python 3.8.

### Ethics

The panel study, PTSD survey, and the prospective study protocols were approved by the Tel Aviv University Institutional Review Board (0007545-1). Before participating in the panel studies, all subjects were advised, in writing, as to the nature of the study. All data were de- identified and no personally identifiable information was gathered.

### Data availability statements

According to this study’s Tel Aviv University Institutional Review Board approval (0007545- 1) and data utilization committees’ guidelines, no patient-level data is to be shared outside the permitted researchers.

### Code availability statements

The Statistical analysis code will be available upon publication.

## Appendix A – Study protocol

### Prospective part

#### Study Design

In this study we will analyze data that were already collected and will be collected as part of the PerMed study ^1^. Participants in the PerMed study are recruited for a period of two years, during which they are equipped with a Garmin Vivosmart 4 smartwatches and are asked to wear them as much as they could. In addition, participants install two applications on their mobile phones: an application that passively collects data from the smartwatch and a dedicated mobile application which allows participants to fill a daily. We will also ask active participants to fill in dedicated surveys for the detection of Post-Traumatic Stress Disorder (PTSD) and anxiety. In this study, we will consider for each participant the 4-weeks period prior to October 7, 2023, as the baseline period.

#### Participants

The inclusion criteria for the PerMed study includes those aged > 18 years. Individuals who are not eligible to give and sign a consent form of their free are excluded. In this study, we will analyze the data of participants aged 18 years and above before and after October, 7, 2023 . To recruit participants and ensure they complete all the study’s requirements, we will hire a professional survey company. Potential participants will be recruited through advertisements in social media, online banners, and word-of-mouth. The survey company is responsible for guaranteeing the participants meet the study’s requirements, in particular, that the questionnaires are filled daily, ensuring the smartwatches are charged constantly and worn properly, and assisting participants resolve technical problems.

#### Study procedures

Before participation in the study, all participants will be advised orally and in writing about the nature of the experiments and give written, informed consent. At this time, participants will be asked to complete an enrollment questionnaire that includes demographic information and health status. In addition, participants will be asked to install two applications on their mobile phones: an application that passively collects data from the smartwatch and the PerMed application, which allows participants to fill in the daily questionnaires. Participants will be given instructions regarding the self-reported symptoms questionnaires and how to operate the smartwatch, which they will wear as much as they can. We will also ask active participants to fill in dedicated surveys of PTSD eight weeks after the events of October 7, 2023 to detect a possible signs of probable PTSD.

#### Enrollment questionnaire

All participants will fill in a one-time enrollment questionnaire that includes demographic questions and questions about the participant’s health condition in general. Specifically, the questionnaire will include the following: age, sex, height, weight, underlying medical conditions, and household income (Listed in Table S2). Other questions such as name, address, phone and email will be recorded and used by the survey company to contact the participants. The answers will be filled-in directly by the survey company to the study’s secured dashboard.

#### Monitoring device

Participants will be equipped with Garmin Vivosmart 4 smart fitness trackers. Among other features, the smartwatch provides all-day heart rate and heart rate variability and during-night blood oxygen saturation level tracking capabilities ^2^.

The optical wrist heart rate (HR) monitor of the smartwatch is designed to continuously monitor a user’s heart rate. The frequency at which heart rate is measured varies and may depend on the level of activity of the user: when the user starts an activity, the optical HR monitor’s measurement frequency increases.

Since heart rate variability (HRV) is not easily accessible through Garmin’s application programming interface (API), we use Garmin’s stress level instead, which is calculated based on HRV. Specifically, the device uses heart rate data to determine the interval between each heartbeat. The variable length of time between each heartbeat is regulated by the body’s autonomic nervous system. Less variability between beats correlates with higher stress levels, whereas an increase in variability indicates less stress ^3^. A similar relationship between HRV and stress was also seen in ^4,5^.

Examining the data collected in our study, we identified an HR sample roughly every 15 seconds, a stress – based HRV sample every 180 seconds.

While the Garmin smartwatch provides state-of-the-art wrist monitoring, it is not a medical-grade device, and some readings may be inaccurate under certain circumstances, depending on factors such as the fit of the device and the type and intensity of the activity undertaken by a participant ^6–8^.

#### Daily questionnaires

All participants will complete the daily self-reported questionnaire in a dedicated application (the PerMed mobile application). The daily questionnaire we will use includes the following questions:

How is your mood today? • Awful (1)• Bad (2)• OK (3)• Good (4)• Excellent (5)

How would you describe the level of your stress during the last day?• Very Low (1)• Low (2)• Medium (3)• High (4)• Very high (5)

How would you define your last night sleep quality?• Awful (1)• Bad (2)• OK (3)• Good (4)• Excellent (5)

Try to remember how many minutes of sports activity you performed on the last day?

Have you experienced one or more of the following symptoms in the last 24 hours?• My general feeling is good, and I have no symptoms• Heat measured above 37.5• Cough• Sore throat• Runny nose• Headache• Shortness of breath• Muscle aches• Weakness / fatigue• Diarrhea• Nausea / vomiting• Chills• Confusion• Loss of sense of taste / smell• Another symptom.

#### PTSD dedicated survey

All participants will be asked to fill in an online survey that includes well-established PTSD surveys via the PerMed application. The Posttraumatic Stress Disorder Checklist (PCL-5) is a self-report questionnaire designed to assess the 20 symptoms outlined in the DSM-5 criteria for PTSD. The PCL-5 serves various purposes, including screening individuals for PTSD and making provisional diagnoses ^9^. It is widely used for clinical and research purposes; psychometrically, the PCL-5 demonstrates strong internal consistency (α = .94), high test-retest reliability (r = .82), and strong convergent validity (rs = .74 to .85) ^9^. We will utilize the General Anxiety Disorder (GAD) 7 item questionnaire (GAD-7) to identify probable cases of GAD along with measuring anxiety symptom severity. The tool is also widely used as a screening measure of panic, social anxiety, and PTSD. The questionnaire is considered valid, sensitive, and specific for the diagnosis of GAD in the general population ^10^. Will use the Israeli Ministry of Health’s (IMOH) Hebrew translation of these surveys. See Appendix D for the full survey.

#### Data Storage

Data collected from the mobile phone application and from the smartwatches will be stored on a secure server within Tel Aviv University facilities. The server runs a CentOS operating system and is located in the Software Engineering Building at Tel Aviv University. This server is protected behind the university’s firewall and is not connected to external networks. In addition, a secure connection through an SSL protocol and a trusted certificate will be obtained for the transfer of information from the mobile phone application into the secured server.

Access will be restricted to investigators in the study. The information from the mobile application will be stored in a structured manner on the secured server without any explicitly identifying information (name, ID number, email). Each participant will be assigned a coded participant number that will be used to identify the subject in the database. The code with the identified information will be stored in an encrypted form on a separate secured server that only the research manager will have access to. Access to all servers is restricted with username and password.

All (non-digital) questionnaires and signed informed consent documents will be stored in a secured cabinet in Tel Aviv University, to which only the research manager and the principal investigators will have access. No data collected as part of the study will be added to individuals’ medical charts.

#### Data preprocessing

We will perform several preprocessing steps. Concerning the daily questionnaires, in cases where participants will fill in the daily questionnaire more than once on a given day, only the last entry for that day will be considered, as it is reasoned that the last one likely best represented the entire day. Self-reported symptoms that are entered as the free text will be manually categorized.

For participant that completed PTSD survey will also be categorized probable PTSD if the total score of the PCL-5 questionnaire is ≥ 33 and meet the DSM-5 diagnostic rule which requires at least: 1 B item (questions 1-5), 1 C item (questions 6-7), 2 D items (questions 8-14), 2 E items (questions 15-20). In line with the GAD-7 criteria, anxiety level of participant will be determined by the total score: no anxiety (0-4), mild (5-9), moderate (10-14) and severe (≥15). Then we will compute the proportion of participants presenting probable PTSD and proportions of each anxiety level.

#### Data Analysis and inclusion criteria

For a more comprehensive understanding of individual perception, we will examine the reported stress levels throughout the PerMed study period. We will calculate the daily centered moving average and the associated 95% confidence interval with a 7-day window from January 1, 2021, for all active participants. For participants who filled in the PTSD online survey, we will implement the same procedure, stratifying them based on whether they exhibited probable PTSD.

For each participant who will remain active after October 7, 2023, we will assess the reported sleep quality, as well as various physiological indicators: duration of awake time during sleep, step counts, and duration of high stress levels. We will compute a single weighted average value for each of the two periods — a week prior to October 7, 2023 (baseline period) and the week after October 7, 2023. For each indicator, we will utilize paired t-test to examine the difference between the weighted averages during the baseline period and those following October 7, 2023.

The weighted average for each participant will be determined by first averaging the corresponding indicator values separately for workdays and free days (weekends and national holidays). Subsequently, the weighted average of these two values will be calculated, assigning a weight of 5/7 to workdays and 2/7 to free days. The rationale for computing this weighted average is further explained in ^11^. In essence, we identified a weekly rhythm across various indicators, with free days exhibiting different mean daily values than workdays. Given the relatively short duration of the examined time periods and variations in the number of free days (e.g., holidays), we aimed to correct for potential biases. Finally, we will calculate the mean and its corresponding 95% confidence interval for each indicator across the participants.

For participants who filled in the PTSD online survey, we will repeat the above analysis, by stratifying the participants based on whether they exhibited probable PTSD.

For each of the well-being indicators, we will explore the unadjusted associations between the probability of probable PTSD, the weighted average during the baseline period, and the difference in the weighted average between baseline and the week after October 7 periods, while controlling for other explanatory variables (age and sex).

All statistical analyses will be performed using Python 3.8.

#### Potential Risks & Risk management

No specific risks arising from the smartwatches are expected, as the device is already commercialized with no known adverse reactions. The main risk in this study is the leakage of private data which we intend to manage as we describe in the following section.

#### Privacy/Confidentiality

Results from this study will be handled at an aggregated level. Individual data records will remain confidential and will not be published or shared with any third party. Signed and dated informed consent forms, as well as data recording sheets (e.g., case report forms) will be stored in locked cabinets during the study and following its completion. A file containing the personal details of the participants will be coded to help preserve confidentiality and will be separated from all other data collected throughout the study. This file will be kept by the principal investigator. Data will be stored on computers in password-protected files.

The data obtained from the smartwatch used in this study will be linked to a coded participant number. The smartwatch does not include a GPS. The data collected by the PerMed application will arrive directly to PerMed back-end servers and will be stored securely.

### Panel study

#### Study Design

In this study will recruit and analyze data of 2,500 participants representative of the Jewish Israeli population in terms of age, sex, and geographical location. Participants will be asked to fill in online dedicated surveys for the detection of probable PTSD and anxiety. Participants will also be asked to provide basic demographic data such as age, sex, educational background, place of residence, and etc. (see Appendix E)

#### Participants

Inclusion criteria includes individuals age > 18 years from the Jewish Israeli population. To recruit participants for the panel study and ensure they complete all the study’s requirements, we will hire a professional survey company. Potential participants will be recruited through a preregistered panelists’ database via email. The survey company will ensure that the participants are representative of the Israeli population in term of age, sex and geographical location. To incentivize participants to complete all study’s requirements (i.e., complete the PTSD survey), each participant will receive 3 New Israeli Shekels upon completion of study’s requirements.

#### Study procedures

Before participating in the study, all participants will be advised in writing about the nature of the survey (see Appendix E). At this time, participants will be asked to complete the PTSD online survey about coping with the events that began on October 7, that includes demographic information and news consumption behavior, to detect a possible sign of probable PTSD nationwide.

#### Description of the data

Data will be collected in the online survey includes (see the survey questions in Appendix E):

- ► Sociodemographic:

- Sex
- Age
- Household size
- Household income
- Religion
- Place of residence (city name)
- ► History of PTSD and anxiety (binary)
- ► News consumption behavior

- News consumption frequency
- News consumption platforms
- ► PCL-5 and GAD-7 questionnaire

#### Data collection and storage

The data will be extracted and stored on a secure server within Tel Aviv University facilities. The server runs a CentOS operating system and is located in the Software Engineering Building at Tel Aviv University. This server is protected behind the university’s firewall and is not connected to external networks.

Access will be restricted to investigators in the study. The information from the online PTSD survey will be stored in a structured manner on the secured server without any explicitly identifying information (name, ID number, email). Each participant will be assigned a coded participant number that will be used to identify the subject in the database.

#### Data Analysis and inclusion criteria

The questionnaire data will be preprocessed by manually categorizing any self-reported symptom of probable PTSD and anxiety according to the PCL-5 and GAD-7 criteria. Participant will be categorized as probable PTSD if the total score of the PCL-5 questionnaire is ≥ 33 and meet the DSM-5 diagnostic rule which requires at least: 1 B item (questions 1-5), 1 C item (questions 6-7), 2 D items (questions 8-14), 2 E items (questions 15-20). In line with the GAD-7 criteria, anxiety level of participant will be determined by the total scores: no anxiety (0-4), mild (5-9), moderate (10-14) and severe (≥15).

We will analyze the data of participants who had either direct or indirect exposure to the events of October 7. We define direct exposure as participants who were evacuated from their place of residence or indicated that they or their immediate family were injured, killed or abducted. Indirect exposure refers to situations in which participants or their immediate family members did not experience physical harm or the need for evacuation, but their income might have been affected. For each level of exposure, we will evaluate the prevalence estimate of PTSD as the proportions of probable PTSD, we will also stratify the prevalence estimate by the duration of news information consumption during the first week after October 7 and the extent of exposure to gory videos. Additionally, we will examine the rates of each anxiety level.

We will also evaluate the rates of exposure to gory videos on each media platform. Specifically, we will divide the number of participants who were exposed to gory videos on each platform by the number of participants who consume news information using that platform.

The 95% confidence intervals (CIs) will be obtained under the assumption of binomial distribution.

To examine the relation between the probability of probable PTSD and news information consumption behavior, controlling for other explanatory variables (age, sex, educational background, PTSD background, religious level, and socioeconomic level), we will fit a logistic regression model.

Likewise, we will also examine the unadjusted associations between the probability of moderate to severe anxiety, duration of news consumption during the two weeks before filling in the online PTSD survey, and the extent of exposure to gory videos, while controlling for other explanatory variables (age, sex, educational background, anxiety background, religious level, and socioeconomic level).

#### Potential Risks & Risk management

The main risk in this study is the leakage of private data which we intend to manage as we describe in the following section.

#### Privacy/Confidentiality

Results from this study will be handled at an aggregated level. Individual data records will remain confidential and will not be published or shared with any third party.

## Appendix B – Data collection platform and data access

### Architecture

The data collection platform contains several components that interact with each other (see Figure 2):

- **The PerMed application** – This application is installed on each participant’s phone to collect sensors data and the self-reported daily questionnaires. It also handles the smartwatch pairing. The current version of the application supports both Android and iOS devices.
- **The smartwatch** - send the data to the Garmin Connect app on the smartphone, which then sends these data to Garmin’s server.
- **The smartwatch application** – This application (currently Garmin) receives information from the smartwatch via Bluetooth and transmits it to the company’s server. In addition, it provides a convenient interface for displaying the participant’s smartwatch information.
- **The app server** – The webserver handles the database connectivity using REST API pages. It enables the server to authenticate users as they launch the application and write records to the database. A MySQL server stores the sensors’ raw data and the answers to the daily questionnaires. At last, there is a batch processes running on the server that sends app notifications (daily reminder to fill the questionnaire).
- **The dashboard server** - hosts the dashboard pages, which assist in monitoring the quality of the information and controlling the experiment. The dashboard has access to participant information and signals indicating whether questionnaires were completed and the smart watch was worn without seeing its content directly. A batch process is responsible for aggregating raw data for dashboard statistics.
- **The smartwatch server** - A MySQL server stores the smartwatch data. A batch process is responsible for collecting the data from the Garmin server.

**Figure S1.**
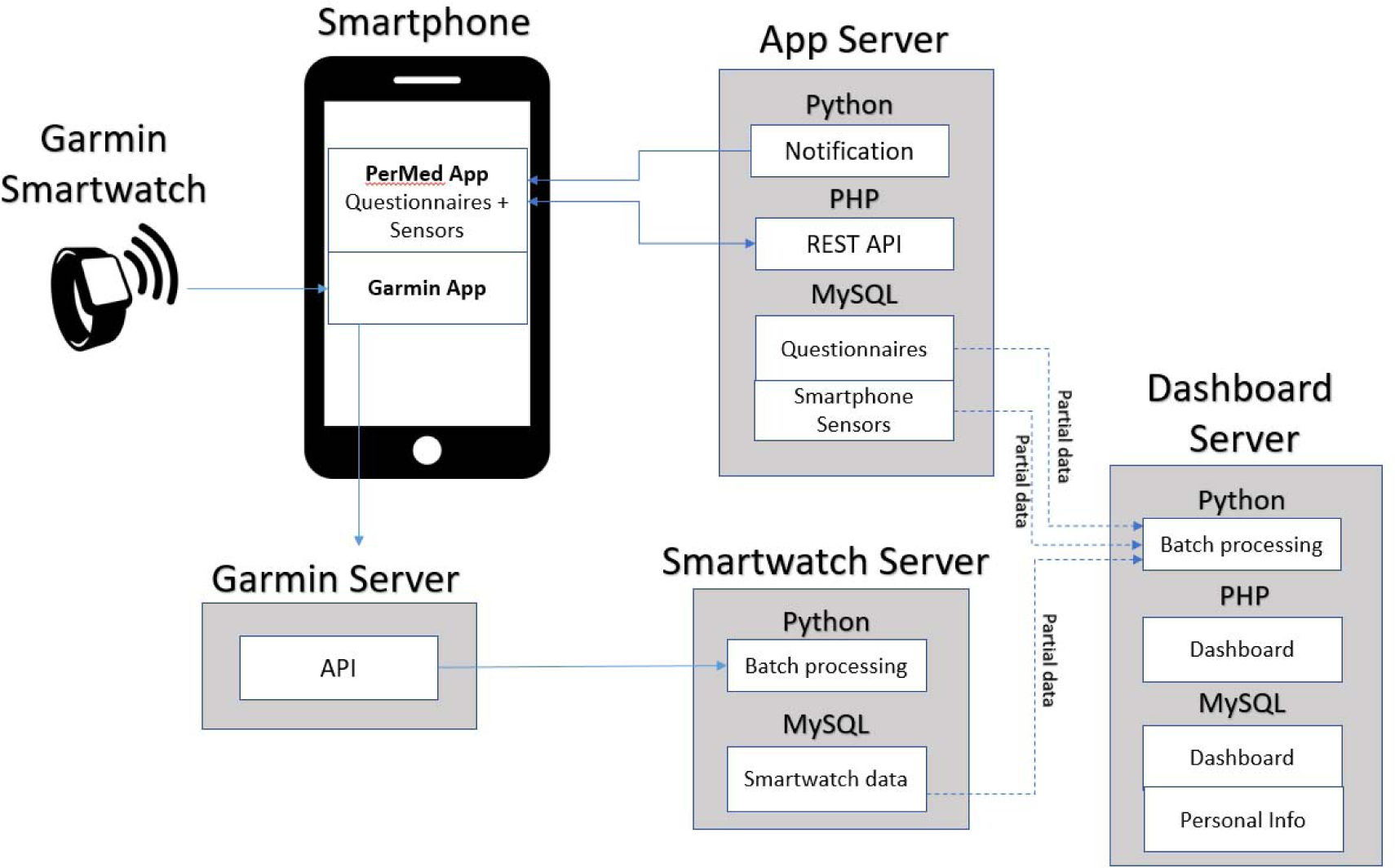
The high-level architecture of the PerMed’s data collection platform.

### The PerMed Dashboard

Participants will be recruited by a qualified external recruitment team headed by Tel Aviv University personal. The team receive limited information essential control the experiment. Thus, we developed a dedicated dashboard for monitoring the quality of the information and control the experiment. This dashboard aims to identify data collection issues such as participants who did not fill the daily questionnaires or participants who did not charge the battery of their smartwatches. The dashboard also helps us identify problems that were not related to participants’ cooperation, such as bugs in the mobileapp. This identification allows us to respond faster and provide timely solutions.

### The Type of Data Collected and data access

Data collected by the platform arrive from four primary sources:

- **Enrolment questionnaire -** data were collected from a one-time enrollment questionnaire that includes basic personal characteristics such as socio-demographic information (e.g., age, gender, height, weight), general habits, health status, and a short Big Five personality questionnaire.
- **Daily questionnaire** – consists of questions on 1) wellbeing, 2) general health condition, 3) symptoms observed, 4) test results to diagnose infectious diseases, 4) vaccination or medication consumption (if relevant to the study question).
- **Smartphone sensor data** – consist of location, Wi-Fi, Bluetooth, screen, and activity.
- **Smartwatch data** - consist of heart rate data, accelerometer and gyroscope information and measures based on these data including active minutes, steps, distance, calories, and sleep level classification, including light, deep, REM, and awake periods.

The current research, aims to explore the safety of vaccination, is part of a larger study. Raw accelerometer data, mobile activity and GPS locations are generally considered sensitive information. In accordance with the data minimization principle, we did not extract this type of data for this vaccination safety research.

## Appendix C – Prospective study participants’ adherence

We employed a professional survey company to recruit participants and ensure they adhere to the study requirements. Participant recruitment was performed via advertisements on social media and word-of-mouth. Each participant signed an informed consent form after receiving a comprehensive explanation on the study. Then, participants completed a one-time enrollment questionnaire, were equipped with Garmin Vivosmart 4 smartwatches, and installed two applications on their mobile phones: (1) the PerMed application ^1,12,13^, which collects daily self-reported questionnaires, and (2) an application that passively records smartwatch data. Participants were asked to wear their smartwatches as much as possible. The survey company ensured that participants’ questionnaires were filled at least twice a week, that their smartwatches were charged and properly worn, and that any technical problems with the mobile applications or smartwatch were resolved. Participants were monitored through the mobile application and smartwatches for a period of at least 49 days, starting seven days before vaccination. Participants also granted full access to their EMR data.

We implemented several preventive measures to minimize participant attrition and discomfort as a means to improve the quality, continuity and reliability of the collected data. First, each day, participants who did not fill their daily questionnaire by 7 pm received a reminder notification through the PerMed application. Second, we developed a dedicated dashboard that allowed the survey company to identify participants who repeatedly neglected to complete the daily questionnaire or did not wear their smartwatch for extended periods of time; these participants were contacted by the survey company (either by text message or phone call) and encouraged to better adhere to the study protocol. Third, to strengthen participants’ engagement, a weekly personalized summary report was generated for each participant, which was available inside the PerMed application. Similarly, a monthly newsletter with recent findings from the study and useful tips regarding the smartwatch’s capabilities was sent to the participants. At the end of the study, participants will receive all personal insights that were obtained and can keep the smartwatch as a gift.

## Appendix D – PTSD online survey

Participants will be asked to fill in an online survey that includes well-established PTSD surveys. The online survey incorporates The Post-traumatic Stress Disorder Checklist (PCL-5) is a self-report questionnaire designed to assess the 20 symptoms outlined in the DSM-5 criteria for PTSD. The PCL-5 serves various purposes, including screening individuals for PTSD and making provisional diagnoses ^9^. It is widely used for clinical and research purposes; psychometrically, the PCL-5 demonstrates strong internal consistency (α = .94), high test-retest reliability (r = .82), and strong convergent validity (rs = .74 to .85) ^9^. We will also utilize the General Anxiety Disorder (GAD) 7 item questionnaire (GAD-7) to identify probable cases of GAD along with measuring anxiety symptom severity. The tool is also widely used as a screening measure of panic, social anxiety, and PTSD. The questionnaire is considered valid, sensitive, and specific for the diagnosis of GAD in the general population ^10^. Will use the Israeli Ministry of Health’s (IMOH) Hebrew translation of these surveys.

The online survey also includes questions regarding demographic information and consumption of news information.

PTSD diagnosis requires an interview conducted by a trained clinician. As our outcome was derived from a screening tool, we characterize the result as a “probable PTSD.” This terminology, utilized in previous studies. Thus, we will opt for a more conservative interpretation, defining only individuals with a total score of ≥ 33 on the PCL-5 questionnaire and meeting the DSM-5 diagnostic rule which requires at least: 1 B item (questions 1-5), 1 C item (questions 6-7), 2 D items (questions 8-14), 2 E items (questions 15-20) as probable PTSD.

**The English version of the PTSD online survey:**

Dear participant,

The following questionnaire is about coping with the events that began on October 7.

The data will be used for statistical analysis as part of an academic study carried out by researchers from the Faculty of Medicine and the Faculty of Engineering from Tel Aviv University.

Your answers will help us learn about the effect of the “Swords of Iron” operation on the mental state of the citizens of the State of Israel and develop ways of coping with the effects of the war.

There are no correct or incorrect answers, but it is crucial to answer all questions truthfully.

The purpose of this questionnaire is not to diagnose any medical condition. If you are feeling unwell, we advise you to contact your primary care physician for further treatment.

Thank you for your valuable time and cooperation with the research team.

**Table S1.**
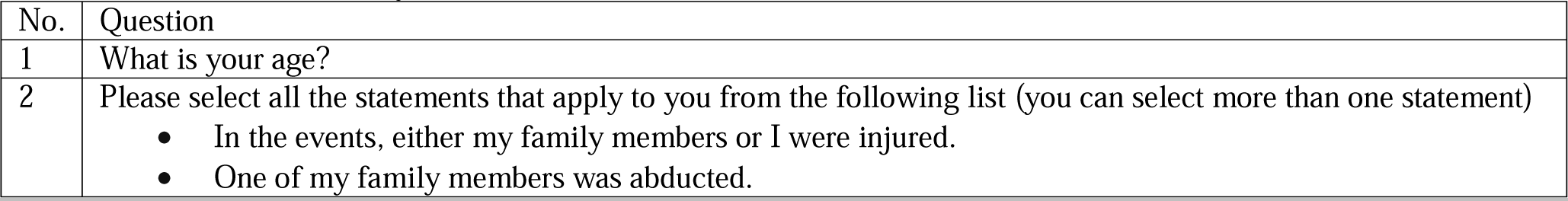
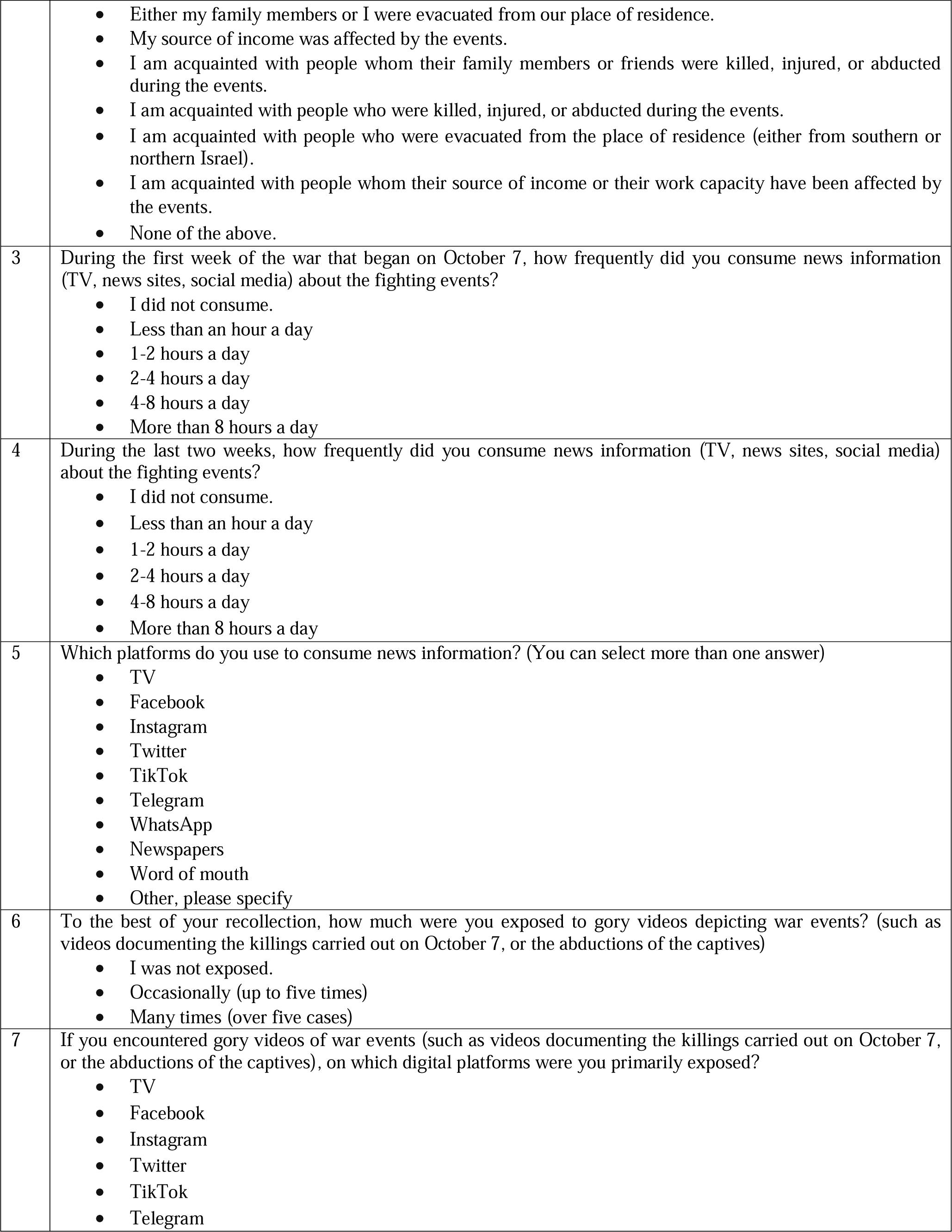
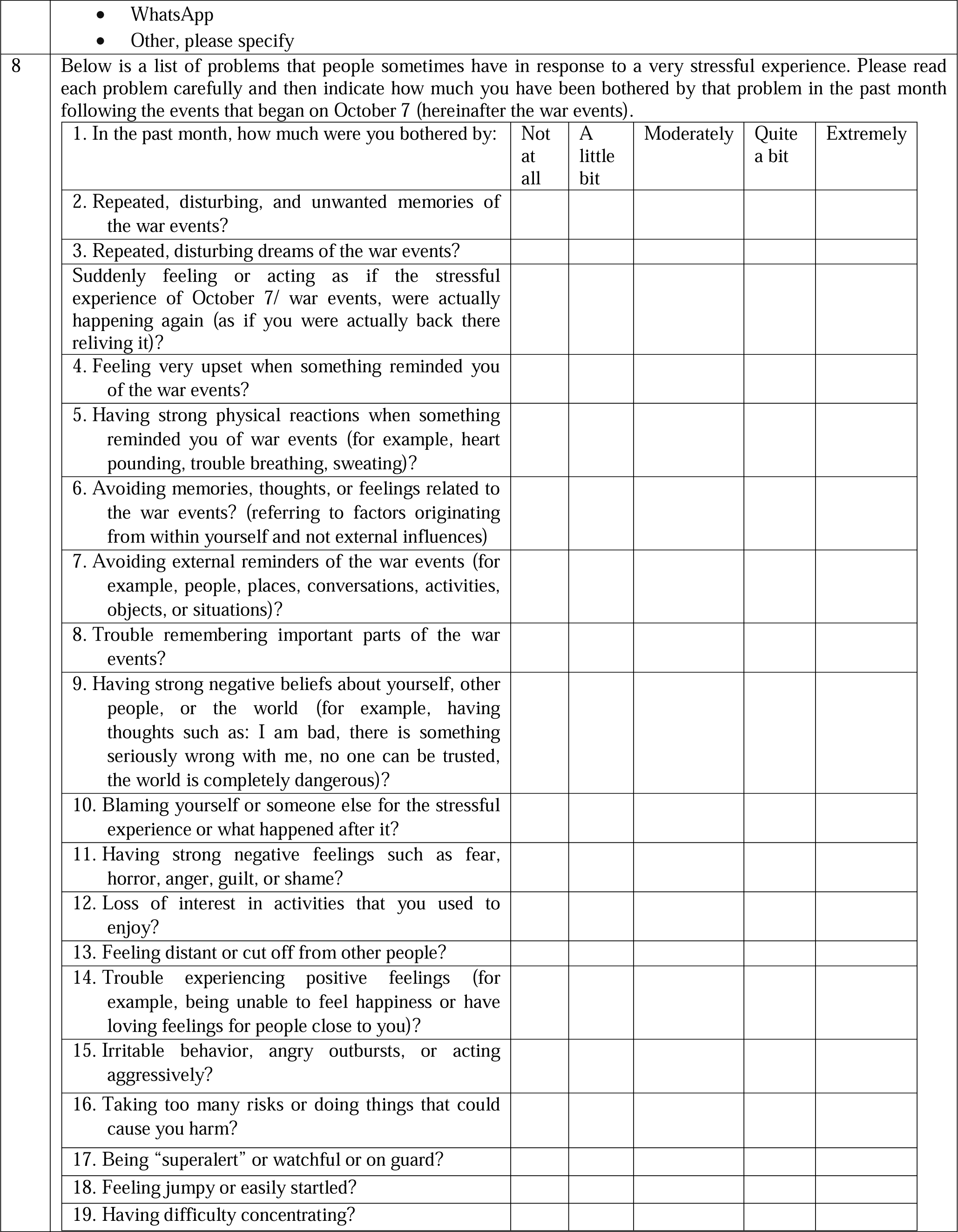
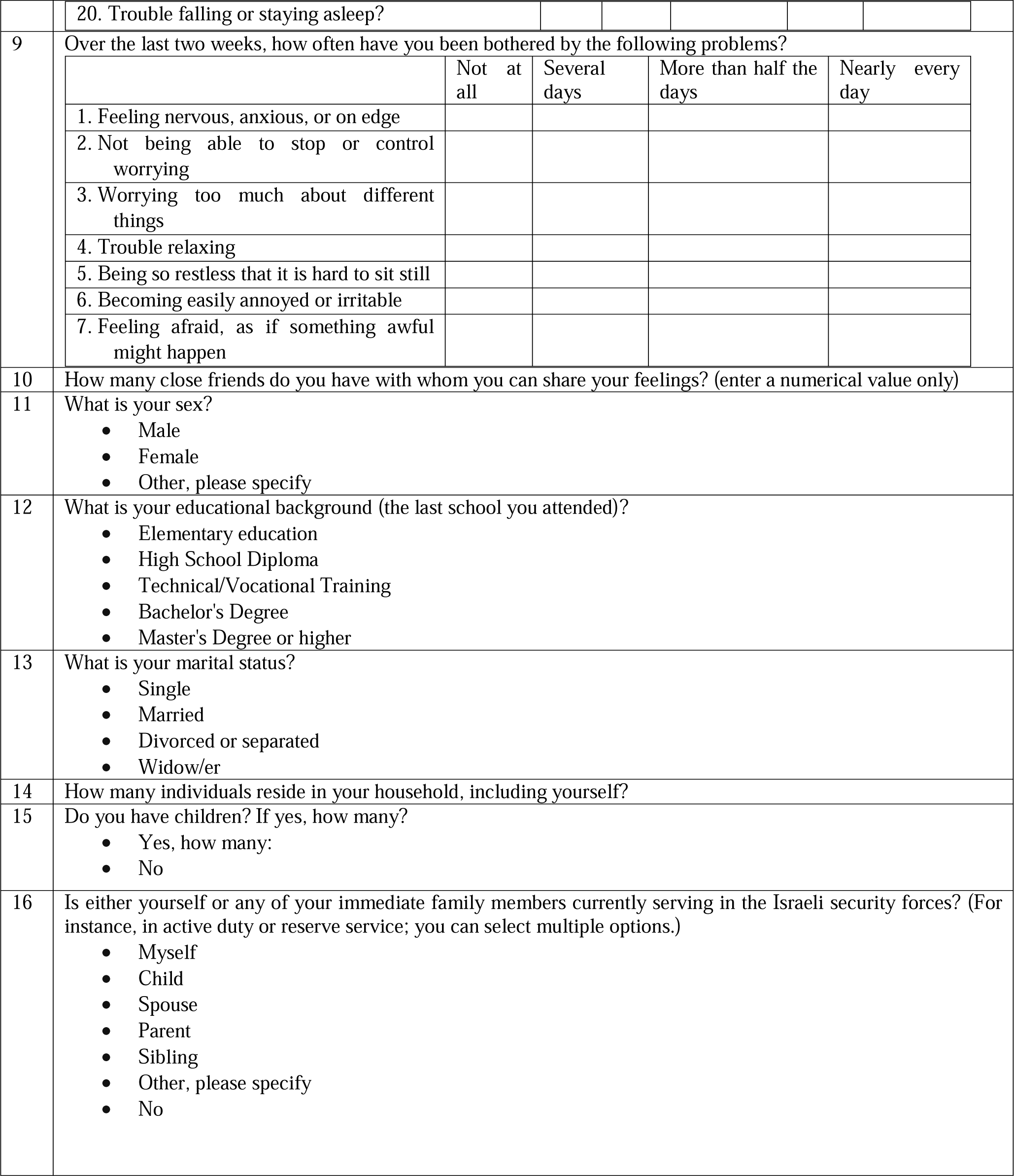
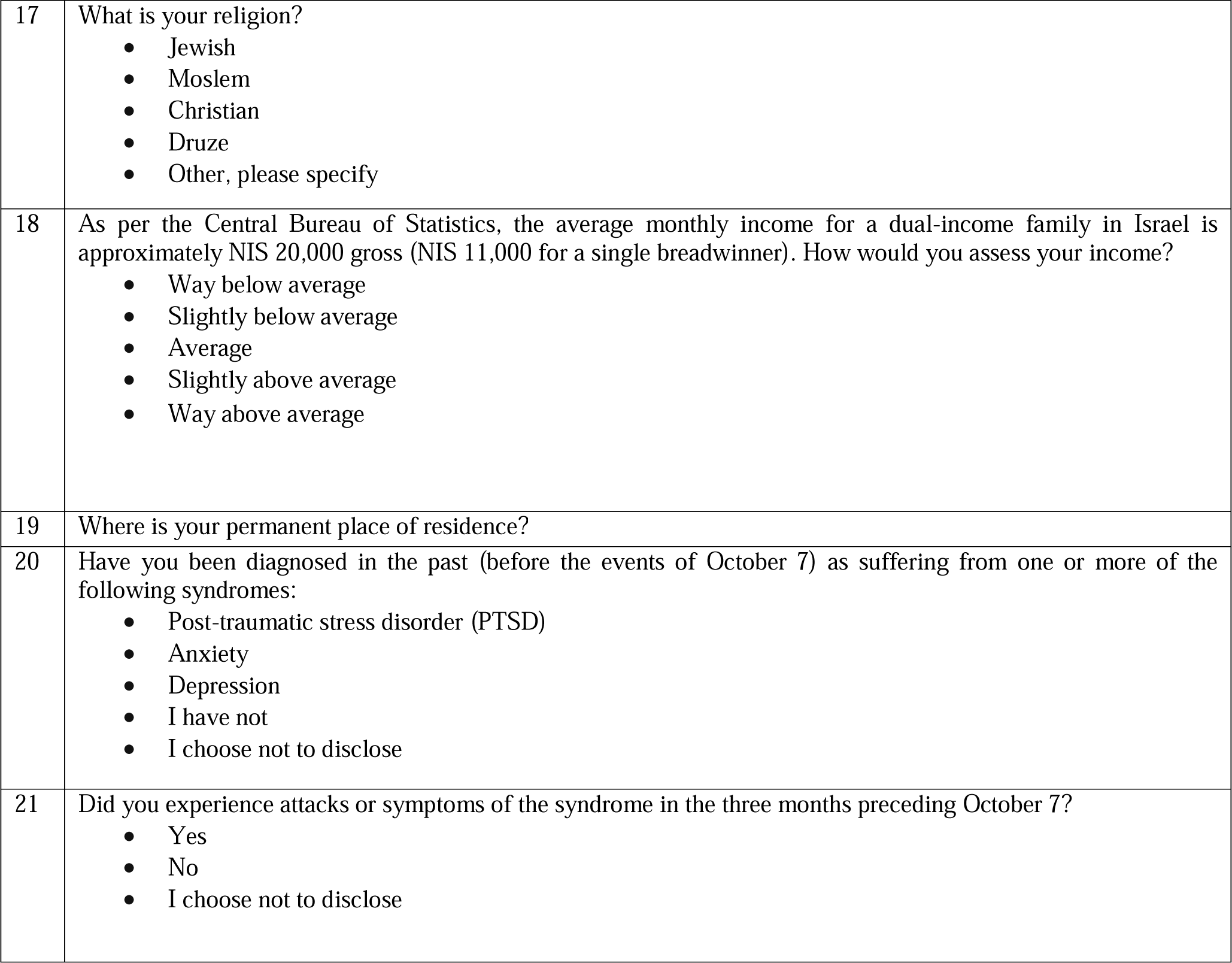
PTSD online survey.

## Appendix E – Additional results

**Table S2.**
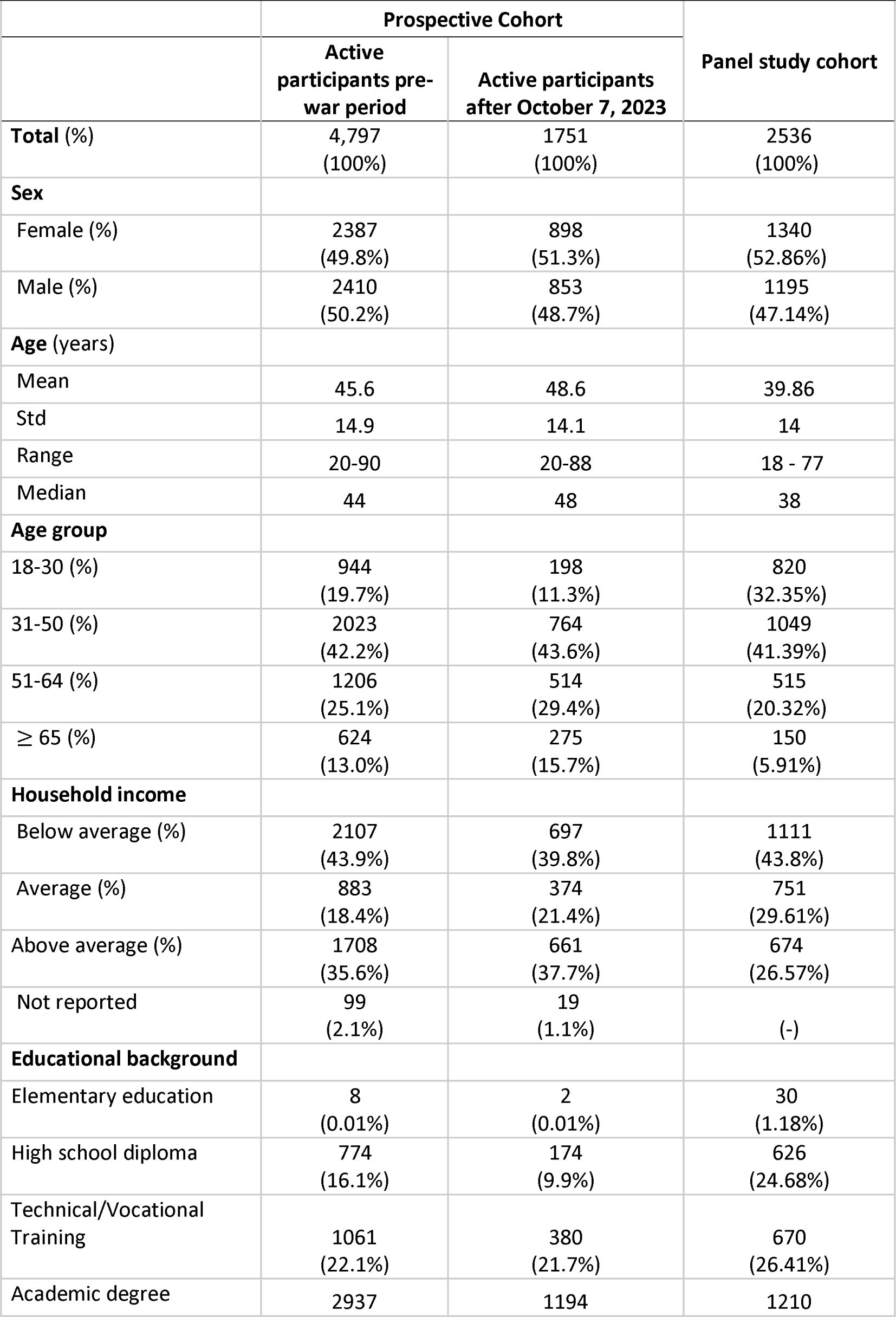
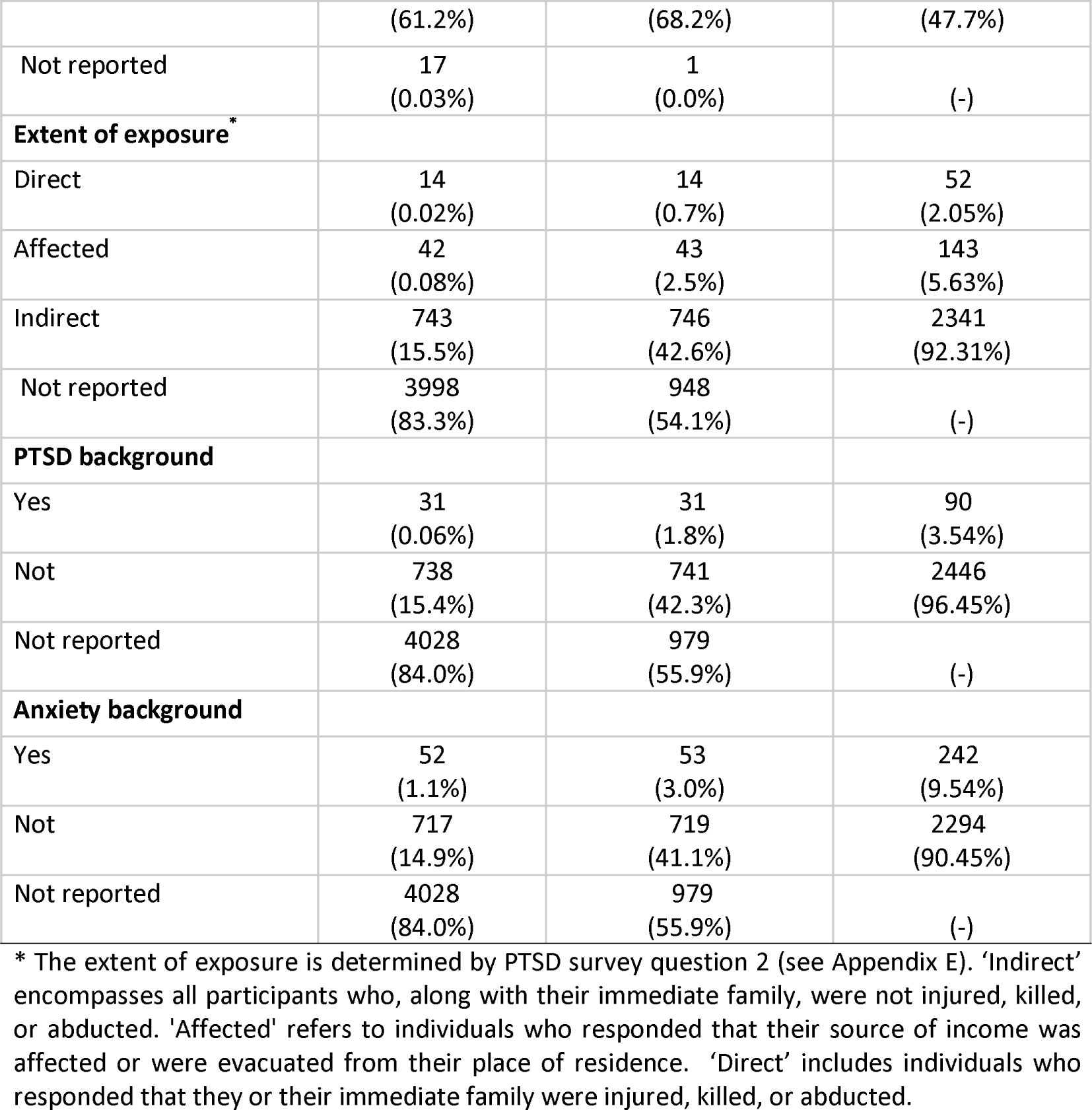
Cohorts characteristics.

**Table S3.** Regression model coefficients for probable PTSD within the participants of the panel study.

| **Factor** | **Coefficient (95% CI)** | **P-value** | **Odds Ratio (95% CI)** |
| --- | --- | --- | --- |
| Age | -0.0195 (-0.028 to -0.011) | <0.001 | 0.981 (0.973 to 0.989) |
| Sex | 0.7462 (0.525 to 0.967) | <0.001 | 2.109 (1.691 to 2.630) |
| Educational background | 0.0833 (-0.025 to 0.192) | 0.132 | 1.087 (0.975 to 1.211) |
| PTSD background | 1.2584 (0.744 to 1.773) | <0.001 | 3.520 (2.104 to 5.887) |
| Duration of news consumption during the first week following October 7 | 0.0845 (0.004 to 0.165) | 0.041 | 1.089 (1.004 to 1.180) |
| Extent of exposure to gory videos | 0.2418 (0.08 to 0.403) | 0.003 | 1.270 (1.083 to 1.497) |
| Religious level | -0.1385 (-0.258 to -0.019) | 0.023 | 0.871 (0.772 to 0.981) |
| Socioeconomic level | -0.1017 (-0.194 to -0.009) | 0.031 | 0.900 (0.823 to 0.991) |

**Table S4.**
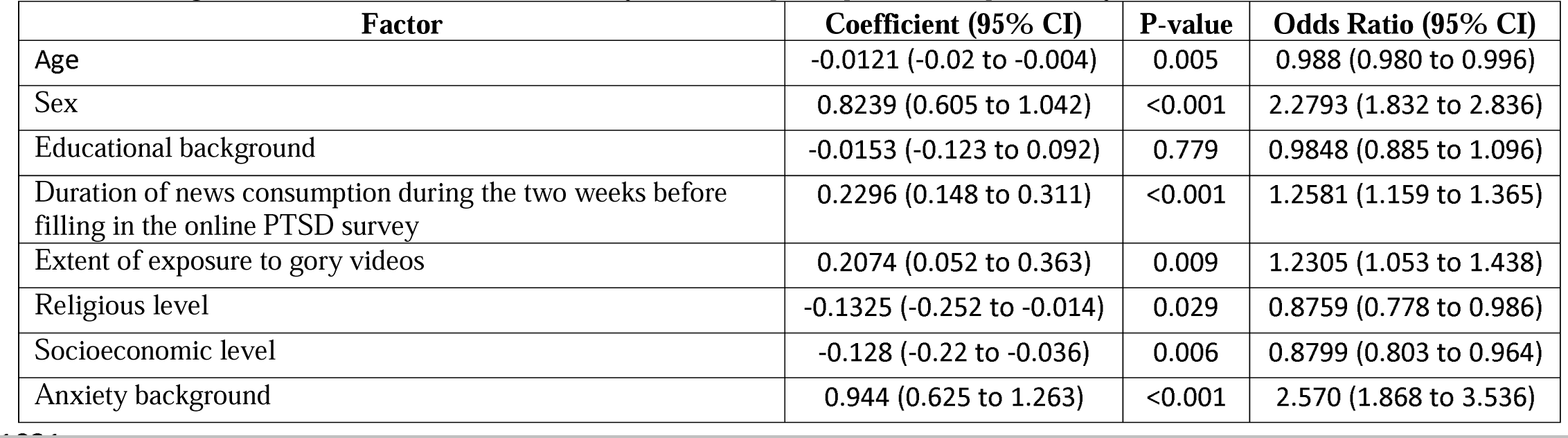
Regression model coefficients for anxiety within the participants of the panel study.

| **Factor** | **Coefficient (95% CI)** | **P-value** | **Odds Ratio (95% CI)** |
| --- | --- | --- | --- |
| Age | -0.0121 (-0.02 to -0.004) | 0.005 | 0.988 (0.980 to 0.996) |
| Sex | 0.8239 (0.605 to 1.042) | <0.001 | 2.2793 (1.832 to 2.836) |
| Educational background | -0.0153 (-0.123 to 0.092) | 0.779 | 0.9848 (0.885 to 1.096) |
| Duration of news consumption during the two weeks before filling in the online PTSD survey | 0.2296 (0.148 to 0.311) | <0.001 | 1.2581 (1.159 to 1.365) |
| Extent of exposure to gory videos | 0.2074 (0.052 to 0.363) | 0.009 | 1.2305 (1.053 to 1.438) |
| Religious level | -0.1325 (-0.252 to -0.014) | 0.029 | 0.8759 (0.778 to 0.986) |
| Socioeconomic level | -0.128 (-0.22 to -0.036) | 0.006 | 0.8799 (0.803 to 0.964) |
| Anxiety background | 0.944 (0.625 to 1.263) | <0.001 | 2.570 (1.868 to 3.536) |

**Table S5.** Paired t-test results for each well-being indicator among the participants of the prospective study.

| Well-being indicator | N | Baseline average value (95% CI) | Week after October 7, 2023, average value (95% CI) | Average difference | P-value |
| --- | --- | --- | --- | --- | --- |
| Reported mood | 228 | 3.68 (3.59 to 3.77) | 2.81 (2.69 to 2.93) | -0.87 | <0.001 |
| Reported stress | 228 | 2.36 (2.26 to 2.46) | 3.28 (3.15 to 3.40) | 0.90 | <0.001 |
| Step counts | 621 | 7881.74 (7588.00 to 8175.47) | 6126.76 (5841.64 to 6411.89) | -1754.97 | <0.001 |
| Distance travelled in meters | 621 | 5983.14 (5675.98 to 6290.29) | 4605.25 (4325.89 to 4884.62) | -1377.89 | <0.001 |
| Reported sleep quality | 227 | 3.51 (3.41 to 3.60) | 3.14 (3.03 to 3.25) | -0.37 | <0.001 |
| Reported sleep time | 225 | 6.61 (6.49 to 6.74) | 6.38 (6.23 to 6.53) | -0.23 | 0.016 |
| Awake duration in seconds | 494 | 594.25 (544.84 to 643.66) | 687.35 (628.82 to 745.88) | 93.10 | 0.017 |
| Light sleep duration in seconds | 494 | 16031.23 (15767.75 to 16294.70) | 16312.04 (16008.71 to 16615.38) | 280.82 | 0.171 |
| Rem sleep in seconds | 494 | 4631.35 (4445.45 to 4817.24) | 4786.15 (4577.13 to 4995.16) | 154.80 | 0.278 |
| Deep sleep duration in seconds | 494 | 4313.71 (4116.09 to 4511.34) | 4304.18 (4085.71 to 4522.65) | -9.54 | 0.949 |

**Table S6.** Regression model coefficients of each well-being indicator among the participants of the prospective study.

| Well-being indicator | Probable PTSD* | N | Baseline average value | Week after October 7, 2023, average value | Average difference | P-value |
| --- | --- | --- | --- | --- | --- | --- |
| Reported mood | No | 187 | 3.71 (3.66 to 3.76) | 2.89 (2.83 to 2.96) | -0.82 | 0.007 |
| Reported mood | Yes | 187 | 3.55 (3.50 to 3.61) | 2.27 (2.20 to 2.34) | -1.28 | 0.007 |
| Reported stress | No | 187 | 2.29 (2.23 to 2.34) | 3.14 (3.07 to 3.21) | 0.85 | 0.002 |
| Reported stress | Yes | 187 | 2.67 (2.60 to 2.73) | 3.98 (3.92 to 4.04) | 1.31 | 0.002 |
| Step counts | No | 431 | 8021.87 (7844.06 to 8199.67) | 6402.28 (6223.12 to 6581.44) | -1619.59 | 0.055 |
| Step counts | Yes | 431 | 7491.50 (7319.69 to 7663.31) | 5336.39 (5188.88 to 5483.90) | -2155.11 | 0.055 |
| Distance travelled in meters | No | 431 | 6258.60 (6056.52 to 6460.67) | 4925.72 (4733.44 to 5117.99) | -1332.88 | 0.079 |
| Distance travelled in meters | Yes | 431 | 5406.74 (5248.06 to 5565.42) | 3783.53 (3667.25 to 3899.81) | -1623.21 | 0.079 |
| Reported sleep quality | No | 186 | 3.58 (3.53 to 3.63) | 3.27 (3.21 to 3.32) | -0.31 | 0.002 |
| Reported sleep quality | Yes | 186 | 3.37 (3.31 to 3.43) | 2.68 (2.62 to 2.74) | -0.70 | 0.002 |
| Reported sleep time | No | 186 | 6.66 (6.59 to 6.72) | 6.48 (6.41 to 6.55) | -0.17 | 0.029 |
| Reported sleep time | Yes | 186 | 6.35 (6.27 to 6.43) | 5.84 (5.72 to 5.96) | -0.51 | 0.029 |
| Awake duration in seconds | No | 357 | 594.21 (565.00 to 623.42) | 665.13 (629.72 to 700.53) | 70.92 | 0.357 |
| Awake duration in seconds | Yes | 357 | 574.45 (551.08 to 597.82) | 738.05 (709.66 to 766.45) | 163.60 | 0.357 |
| Light sleep duration in seconds | No | 357 | 15911.31 (15760.01 to 16062.60) | 16079.81 (15904.66 to 16254.95) | 168.50 | 0.371 |
| Light sleep duration in seconds | Yes | 357 | 16797.03 (16645.68 to 16948.39) | 17137.82 (16967.12 to 17308.52) | 340.79 | 0.371 |
| Rem sleep in seconds | No | 357 | 4730.07 (4612.17 to 4847.97) | 4781.87 (4655.31 to 4908.43) | 51.80 | 0.810 |
| Rem sleep in seconds | Yes | 357 | 4559.48 (4461.32 to 4657.63) | 4661.21 (4549.23 to 4773.19) | 101.74 | 0.810 |
| Deep sleep duration in seconds | No | 357 | 4281.51 (4161.61 to 4401.40) | 4343.37 (4209.04 to 4477.69) | 61.86 | 0.600 |
| Deep sleep duration in seconds | Yes | 357 | 4344.85 (4237.13 to 4452.57) | 4296.20 (4164.64 to 4427.75) | -48.65 | 0.600 |

\* Probable PTSD was determined based on the response for the PTSD survey (see Appendix D)

## Appendix F – Additional results: continous physiological measures

To calculate the changes in smartwatch measures after October 7, 2023, for active participants in the prospective study, (Figures S2 and S3), we performed the following steps. First, for each participant and each hour we calculated the difference between the mean value of the measure tested on that hour and that of the corresponding hour in a week prior (keeping the same day of the week and same hour during the day). If such data were not recorded (e.g., a participant did not wear the smartwatch in the same period before and after vaccination), we excluded the participant from this analysis. Then, we aggregated each hour’s differences over all participants to calculate a mean difference and the associated 95% confidence interval.

**Figure S2.**
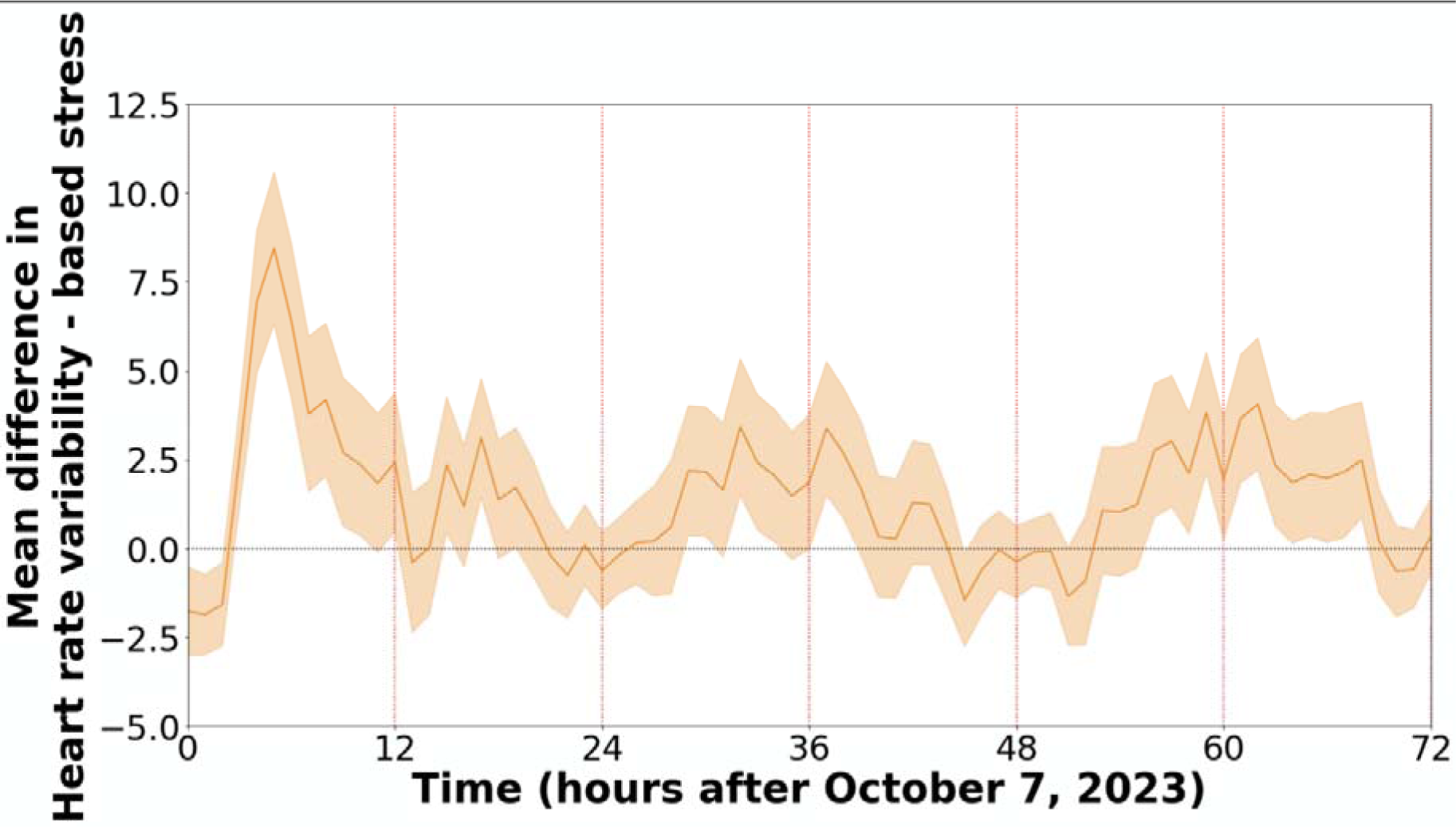
HRV-based stress reaction to October 7, 2023, events as recorded by the smartwatches. The figures show the mean difference between the baseline and the week after period in terms heart rate variability-based stress (n=512). Mean values are depicted as solid lines, and 95% confidence intervals are presented as shaded regions.

**Figure S3.**
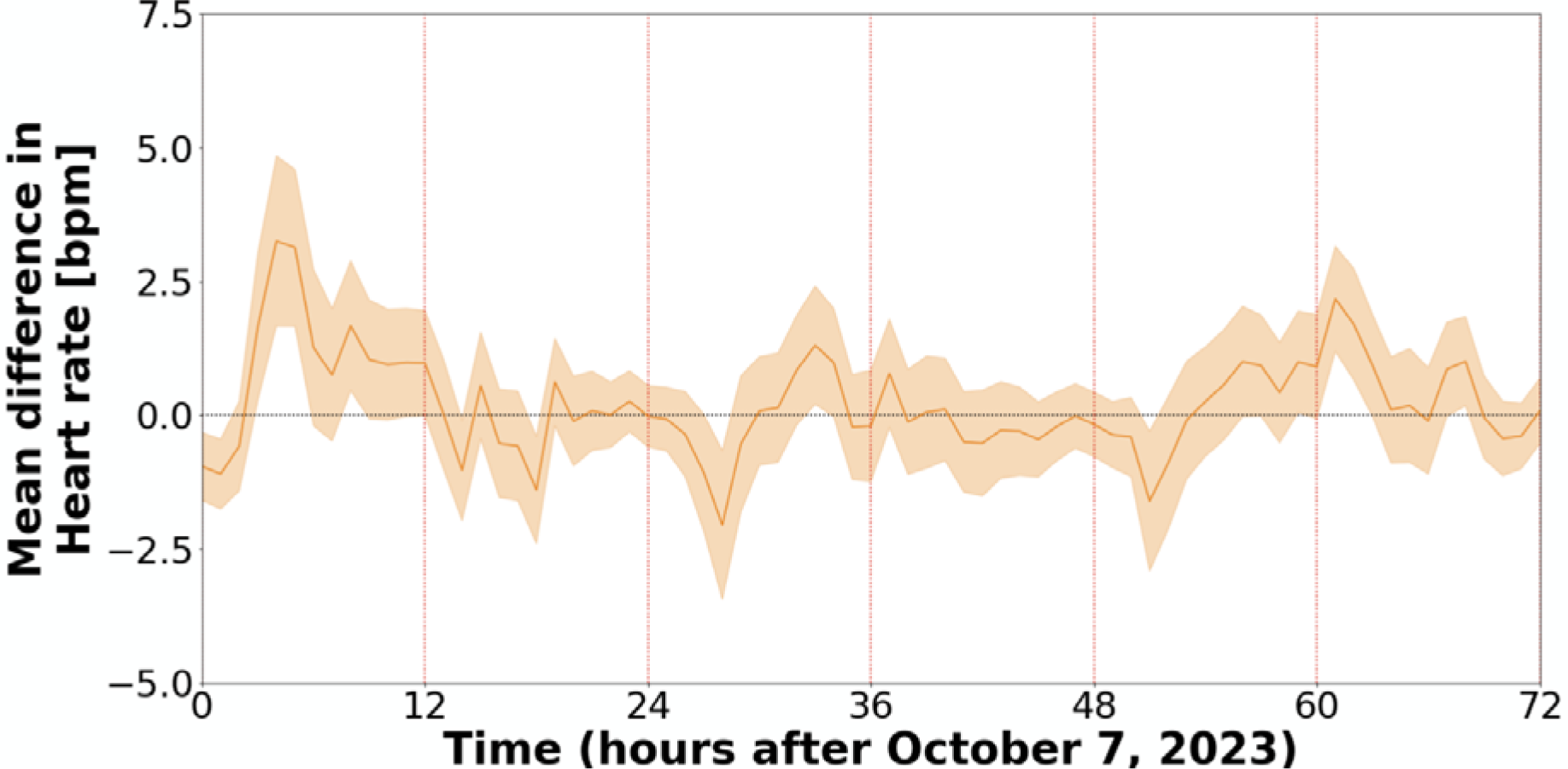
Heart rate reaction to October 7, 2023, events as recorded by the smartwatches. The figures show the mean difference between the baseline and the week after period in terms heart rate (n=495). Mean values are depicted as solid lines, and 95% confidence intervals are presented as shaded regions.

